# Complementarity of long-read sequencing and optical genome mapping in Parkinson’s disease

**DOI:** 10.1101/2025.08.20.25333965

**Authors:** André Fienemann, Theresa Lüth, Susen Schaake, Carolin Gabbert, Marius Möller, Hauke Busch, Katja Lohmann, Jonas A. Gustafson, Danny E. Miller, Kensuke Daida, Manabu Funayama, Nobutaka Hattori, Samia Ben Sassi, Faycel Hentati, Matthew J. Farrer, Kristian K. Ullrich, Christine Klein, Joanne Trinh

## Abstract

**Background:** With third-generation long-read sequencing (LRS) platforms and optical genome mapping technologies (OGM), the ability to detect large and complex structural variants (SVs) is rapidly advancing. This has led to the discovery of novel pathogenic variants, such as large deletions and insertions, in neurodegenerative movement disorders. Thus, we aimed to systematically examine the applicability of the combined application of LRS and OGM in Parkinson’s disease (PD).

**Methods:** Ultra-high molecular weight DNA was derived from blood and fibroblast cultures and used for Oxford Nanopore Technologies (ONT) LRS and OGM. We included 19 patients with mostly early-onset PD. Variant calling was performed with the tools Sniffles2 and Spectre for ONT and the Bionano Solve software for OGM. The size distribution of deletions and insertions was compared, and a subsequent analysis pipeline based on AnnotSV, SVAFotate, and needLR was employed to annotate and filter for rare (population allele frequency ≤1%) or potentially pathogenic (CADD-SV >20) variants affecting 134 known movement disorder genes.

**Results:** Both methods identified SVs ≥50 kb; however, OGM detected fewer SVs (49,677) with a larger mean size of 25 kb (SD=209 kb) compared to ONT (92,030, mean=17 kb, SD=1.1 Mb). In the size bracket of 50-80 kb, which falls outside the ideal detection range of Sniffles2 and Spectre, OGM detected 384 deletions and insertions, compared to six detected by ONT. OGM detected significantly larger deletions and insertions than ONT (p-value <2.2×10^-16^). Regarding known movement disorder genes, a heterozygous intergenic deletion (195 kb) near *ITPR1* was detected by both methods, and OGM validated a previously published 7 Mb inversion in *PRKN*. Heterozygous deletions in *ATXN2* (1.4 kb), *SUCLA2* (1.7 kb), and *PNKD* (2.6 kb) were detected by OGM and confirmed to be intronic by ONT.

**Conclusion:** OGM allows for better detection of large insertions and can serve as a powerful first-line method to detect large pathogenic variants. However, it greatly benefits from a high-resolution sequencing technique like ONT to refine breakpoint positions. Despite certain limitations, ONT proved to be highly capable of detecting large variants independently; thus, it allows for a highly complementary assessment and validation of structural variation in combination with OGM.

## Introduction

Structural variants (SVs) are one of the largest contributors to genetic variation between human genomes (1, 2) and are commonly defined as alterations that exceed 50 nucleotides in length. Subtypes include deletions, duplications, and insertions of genomic sequence, as well as balanced inversions and translocations, all of which can disrupt genes and regulatory elements, contributing to complex genomic rearrangements with potentially catastrophic consequences. Due to its high base-calling accuracy, short-read exome sequencing is widely applied in genetic research, particularly for detecting pathogenic single-nucleotide variants (SNVs) or short indels (3-5). However, its capabilities for detecting SVs of comparable size to the read length or even larger SVs are limited, especially affecting insertions or complex rearrangements (6-10). In the field of neurodegenerative movement disorders, long-read sequencing (LRS) methods like single-molecule real-time (SMRT) technology by Pacific Biosciences (PacBio) or Nanopore sequencing by Oxford Nanopore Technologies (ONT) have the potential to help fill in remaining gaps in our genetic understanding of these severe diseases (11-13). In Parkinson’s disease (PD) research, ONT was recently performed for *N*=47 affected individuals with recessively inherited forms of PD linked to pathogenic variants in *PRKN* and *PINK1* to identify previously undetected secondary variants (14). Out of twenty-three heterozygous *PRKN* carriers, ONT identified six (∼26.1%) with additional SVs in trans (14). The functional consequence of a non-coding African ancestry-specific *GBA1* risk variant was identified using long-read RNA sequencing despite the nearby *GBA1LP* pseudogene exhibiting high sequence homology (∼96%) (11). In the context of the first short-read-based SV GWAS in PD, further benchmarking using LRS revealed that ∼30% of the identified SVs were false positives, and ∼85% of SVs detected with long-read sequencing could not be detected using short-read sequencing (13, 15). Finally, ONT was able to identify a large 7 Mb inversion disrupting exon 12 of *PRKN* in compound heterozygous affected twins with early-onset PD (16).

Optical genome mapping (OGM) is a different approach to whole-genome sequencing that is more akin to classic cytogenetic tools, such as fluorescence in situ hybridization (FISH) and karyotyping, but greatly surpasses them in terms of resolution and sensitivity (17, 18). It has been used to identify and size repeat expansions in *RFC1* that cause late-onset ataxia (19, 20), demonstrating higher accuracy compared to Southern Blotting (19). Despite its high accuracy, OGM is often applied to validate a detected variant rather than as a first-line technique (21, 22). One limitation of OGM is the lower resolution threshold of 500 bp compared to sequencing methods like ONT, which can capture single-base pair changes. However, molecules imaged during OGM have a very high N50 of ≥ 150kb and are measured to a coverage of ≥ 100X, allowing for high sensitivity and confidence (23). These large fragment sizes also allow for effective phasing, as recently demonstrated for large compound heterozygous *PRKN* deletions (24).

In this study, we compared the SV detection capabilities of ONT and OGM for the human whole genome of patients with PD as a use case. We evaluated their concordance and complementarity genome-wide and for potentially pathogenic variants in 134 known genes associated with movement disorders presented by the Movement Disorder Society (MDS) Taskforce (25) to benchmark their ability to identify different types and sizes of medically relevant SVs.

## Methods

### Research cohort

Nineteen patients affected by idiopathic PD were examined using OGM and ONT LRS (Figure 1). Fourteen were of central European descent, with a subset of five Tunisian individuals (Supplementary Table 1). Preferentially, patients exhibiting an early disease onset (average onset age ±SD: 39±14 years; range, 14–73 years) were included to increase the likelihood of detecting a monogenic disease cause. All patients provided informed consent, and local ethics committee approval was obtained at the University of Lübeck and the University of British Columbia. As a separate case study outside of the cohort, OGM was performed on a single sample of a patient with PD originating from Japan, who carried a known 7 Mb inversion affecting *PRKN* (16).

**Figure 1:**
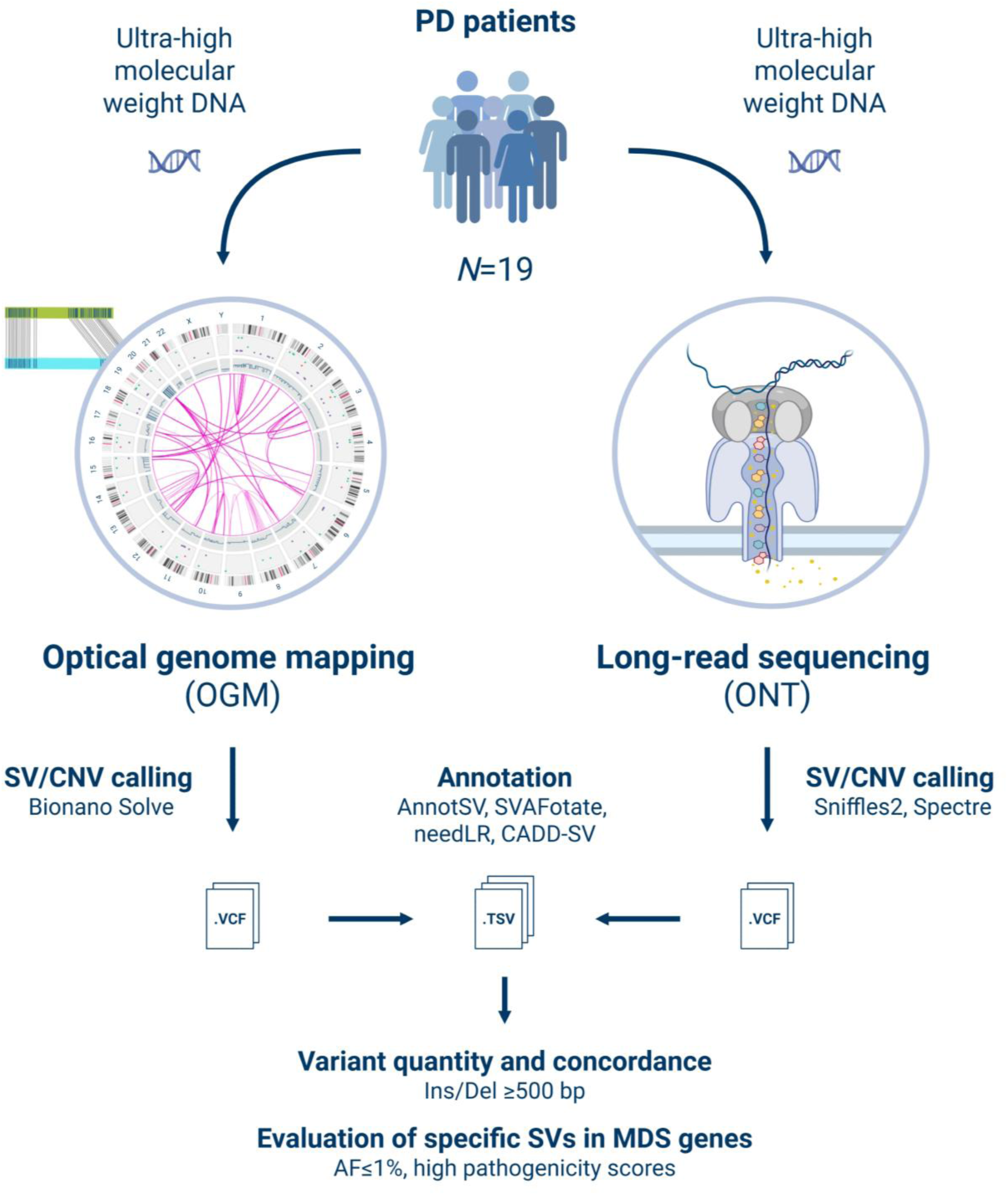
Overview of the analysis workflow. Whole blood and skin biopsies (to grow fibroblast cultures) were obtained from *N*=19 predominantly early-onset PD patients and used to isolate genomic ultra-high molecular weight DNA (UHMW). The SV detection capabilities of ONT and OGM were compared. SV, structural variant; CNV, copy number variation; Ins, insertion; Del, deletion; MDS, Movement Disorder Society; AF, allele frequency. Created in BioRender. Fienemann, A. (2025).

### Optical genome mapping

Whole blood or skin biopsies were obtained from the cohort. Skin biopsies were used to grow fibroblast cultures in Dulbecco’s Modified Eagle’s Medium supplemented with 10% fetal bovine serum and 1% penicillin/streptomycin (Thermo Fisher Scientific). Genomic ultra-high molecular weight DNA (UHMW) was isolated from blood or fibroblast cell pellets with the "Bionano Prep SP-G2 Frozen Human Blood DNA Isolation Protocol" or the "Bionano Prep SP-G2 Frozen Cell Pellet DNA Isolation Protocol", depending on the available biomaterial according to the manufacturer’s instructions. In brief, after lysis of the surrounding tissue, the UHMW DNA was precipitated on a nanobind magnetic disk using isopropanol (≥99.5%) and washed. Afterward, the UHMW DNA was eluted, homogenized, and quantified using Qubit broad-range double-stranded DNA assay kits (Thermo Fisher Scientific). DNA labeling was conducted according to the "Bionano Prep DLS-G2 Protocol". 750 ng of the UHMW DNA were incubated with the DLE-1 enzyme and DL-Green fluorophores, resulting in covalently attached labels at specific sequence motifs. After the proteolytic digest of DLG-1, removal of the remaining fluorophores, and another quantification step, the labeled DNA molecules were loaded on a Saphyr flowcell chip and imaged on the Bionano Saphyr. Following the optical imaging, the generated optical maps were processed with the *de novo* pipeline included in the Bionano Solve software (v3.8). During this process, the images are aligned against each other, assembled, and compared to the GRCh38 reference genome to identify SVs. The subsequent results of the *de novo* pipeline were analyzed via the Bionano Access software (v1.8), and the identified SVs and CNVs were exported as VCF and SMAP files containing the variants.

### Nanopore long-read sequencing

LRS was performed at two different facilities using UHMW DNA extracted for OGM: externally at the NGS Competence Center Tübingen (NCCT) (*N*=7) and internally at the Institute of Neurogenetics (ING) Lübeck (*N*=12). At the NCCT, the SQK-LSK109 (ONT) library kit and R9.4.1 flow cells (FLO-PRO002) were used for library preparation and sequencing on an ONT PromethION device. Pre-treatment with T7 endonuclease was performed prior to library preparation and flow cell loading according to the standard SQK-LSK109 protocol. The base-calling was performed using Dorado v0.6.3. (basecall server 7.4.12).

At the ING, UHMW DNA was sheared with the Megaruptor 3 (Diagenode), and subsequent size selection was performed using the BluePippin (Sage Science). The Qubit fluorometer (Thermo Fisher Scientific), Femto Pulse system, and Tapestation 4200 (Agilent Technologies) were used to quantify the DNA and monitor the size range after shearing and size selection (26). Library preparation and sequencing were conducted using the manufacturer’s SQK-LSK114 protocol for genomic DNA, R10.4.1 flow cells (FLO-PRO114M), and the PromethION device from ONT. High accuracy base-calling (HAC) was performed using Dorado v0.6.3 in MinKNOW v24.06.10, generating output files (.fastq) for in-house analysis.

### Data analysis

The ONT fastq files containing the sequence information were concatenated and converted to an unmapped BAM file using v1.9 of Samtools (27). This input was analyzed using v2.3.1 of the wf-human-variation workflow of EPI2ME Labs (github.com/epi2me-labs/wf-human-variation), a collection of tools supported by ONT. This includes alignment to the GRCh38 reference genome using v2.28 of minimap2 (28) as well as variant calling using v2.3.3 of Sniffles2 for SVs (29, 30) and v0.2.1 of Spectre for CNVs (31). The newer v2.6.2 standalone version of Sniffles2 was additionally applied to selected samples to evaluate the difference in large variant detection for specific variants. The Sniffles2 and Spectre variants, as well as the OGM variants detected by Bionano Solve, were annotated using v3.4 of AnnotSV (32) and v1.0 of SVAFotate (33) to obtain functional and regulatory information as well as population-level allele frequencies based on v4.1.0 of the SV callset of the Genome Aggregation Database (gnomAD SV). To account for larger variants not present in the short-read-based gnomAD SV database, the variants were additionally annotated using v3.4 of needLR (34) to obtain allele frequencies based on the ONT LRS data from the 1KGP-ONT Consortium (450 samples). V1.1.2 of CADD-SV (35) was applied to score the potential effect of coding and especially noncoding SVs. The variants were assessed and visualized using R (v4.5.0) (36) with the packages data.table (v1.17.2) (37), tidyverse (v2.0.0) (38), dplyr (v1.1.4) (39), and ggplot2 (v3.5.2) (40). Only variants with a Phred score of at least 20 (corresponding to 99% confidence and the maximum for OGM variants) or -1 (confidence cannot be assessed) were considered. Wilcoxon signed-rank tests were performed on the length distribution of deletions and insertions detected by both OGM and ONT to test for differences in the detected variant length. To estimate the concordance of both methods, ONT variants ≥500 bp (resolution of OGM) were compared to the OGM variants and matched according to the SV/CNV type, chromosome, position in the GRCh38 reference genome, and variant length. Insertion and duplication calls were combined into one category to account for different nomenclatures between variant callers (Liu et al., 2024). As an approximation, a variant was considered a match if the difference in the SV’s start, end and length did not exceed an interval of ±6 kb for variants ≤50 kb, ±15 kb for variants ≥50 kb or ±7 kb for variants in movement disorder genes, based on the median uncertainty (95% CI) of the OGM variant calls (Supplementary Table 2). Rare variants with a population allele frequency of ≤1% in either gnomAD SV or the internal Bionano database, variants with high pathogenicity prediction, e.g., pathogenic according to the ACMG (41) or CADD-SV≥20, or variants in or near known movement disorder genes presented by the MDS Taskforce (25), were evaluated to compare the performance and concordance of both methods regarding the detection of novel specific variants (Supplementary Figure 1).

## Results

### Molecule and read statistics

For each OGM run, on average 2,289,579 molecules with a mean length of 240.4 kb were imaged. A mean N50 of 237.5 kb and a mean coverage of 176.7 X were obtained. A mean label density of 15.9 labels per 100 kb was measured (Table 1), which is in the expected range of 14–17/100 kb for optical mapping (42). With ONT, a mean of 7,229,440 reads with a mean length of 14.6 kb were sequenced with a mean N50 of 25.8 kb and a mean coverage of 26.8 X. A mean Phred quality score of 15.3 was obtained (Table 1).

**Table 1:** Molecule and read statistics. Overview of different metrics describing the molecules imaged during OGM and the reads sequenced during ONT sequencing of the cohort.

| Bionano optical genome mapping |  |  |  |  |  |  |
| --- | --- | --- | --- | --- | --- | --- |
|  | Total molecules | Total length (Mb) | Mean length (kb) | Molecule N50 (kb) | Label density (x/100 kb) | Coverage (X) |
| **Mean** | 2,289,579 | 545,936 | 240.35 | 237.51 | 15.9 | 176.7 |
| **Median** | 2,160,768 | 512,296 | 239.67 | 235.61 | 15.9 | 165.9 |
| **SD** | 607,168 | 122,522 | 22.11 | 28.22 | 0.5 | 39.7 |
| Oxford Nanopore long-read sequencing |  |  |  |  |  |  |
|  | Total reads | Mean Q-score | Mean length (bp) | Read N50 (kb) |  | Coverage (X) |
| **Mean** | 7,239,301 | 15.3 | 14,587 | 25.80 |  | 26.8 |
| **Median** | 6,065,873 | 14.9 | 16,036 | 31.84 |  | 24.6 |
| **SD** | 3,949,356 | 3.3 | 6,936 | 11.44 |  | 6.3 |

The cohort comprised $N=19$ individuals. bp: base pairs, kb: kilobase, Mb: megabase, Q-score: read quality (Phred). X: depth of coverage.

### Structural variant detection with OGM and ONT

Across all patients, OGM identified 77,119 SVs, corresponding to a mean of 4,056 SVs per individual (Table 2). After filtering for high-confidence variant calls (Confidence ≥99%, Phred quality score ≥20), a total of 49,677 variants remained. A total of 804 high-quality variants showed a rare population allele frequency of ≤1% in the internal Bionano control database. There were 299 high-quality variants in known movement disorder genes, four of which were rare.

**Table 2:**
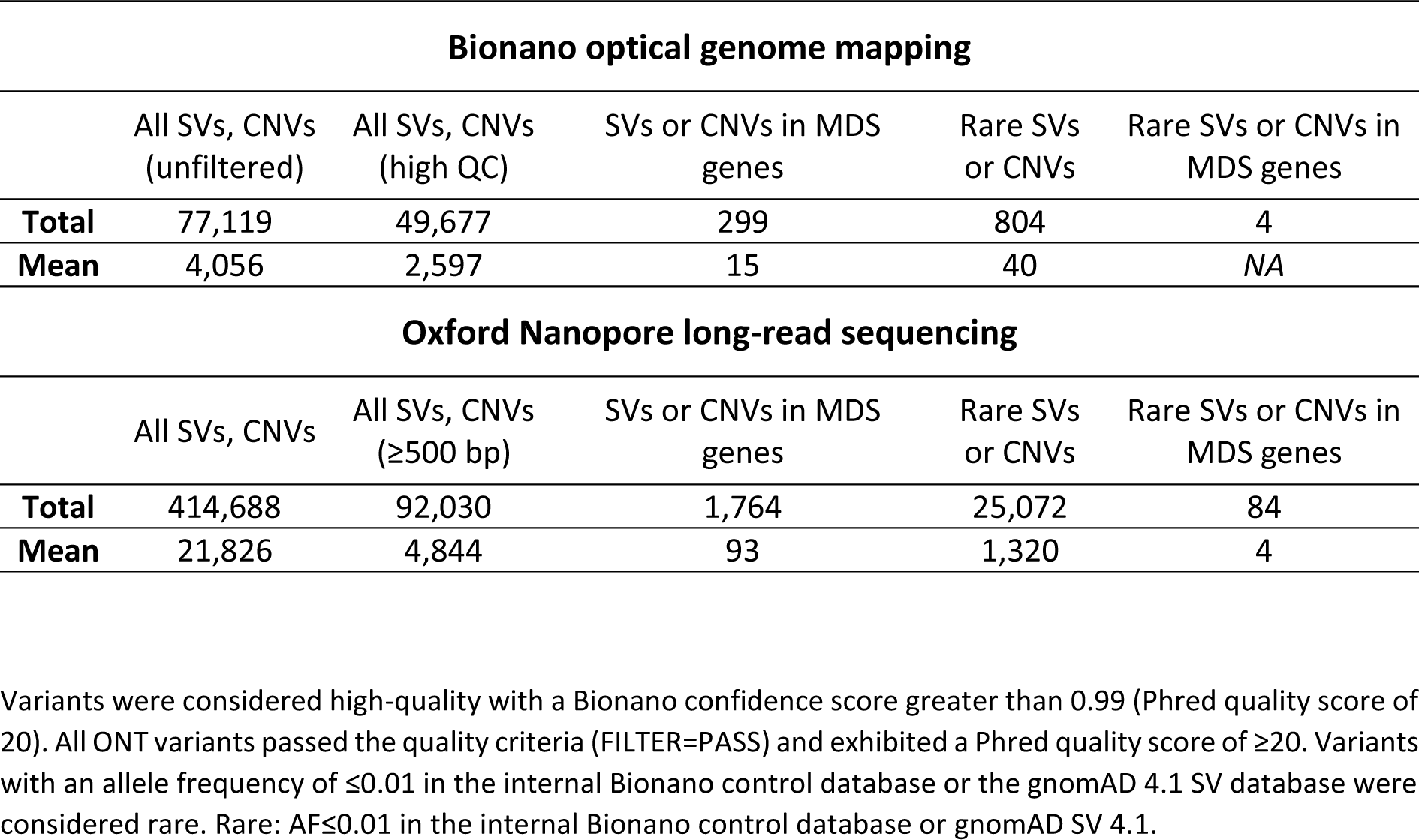
Quantity of SVs and CNVs. High-quality SVs and CNVs identified with OGM and ONT, as well as filtered subsets for rare variants and variants in known genes associated with movement disorders.

| **Bionano optical genome mapping** |  |  |  |  |  |
| --- | --- | --- | --- | --- | --- |
|  | All SVs, CNVs<br>(unfiltered) | All SVs, CNVs<br>(high QC) | SVs or CNVs in MDS<br>genes | Rare SVs<br>or CNVs | Rare SVs or CNVs in<br>MDS genes |
| **Total** | 77,119 | 49,677 | 299 | 804 | 4 |
| **Mean** | 4,056 | 2,597 | 15 | 40 | NA |
| **Oxford Nanopore long-read sequencing** |  |  |  |  |  |
|  | All SVs, CNVs | All SVs, CNVs<br>(≥500 bp) | SVs or CNVs in MDS<br>genes | Rare SVs<br>or CNVs | Rare SVs or CNVs in<br>MDS genes |
| **Total** | 414,688 | 92,030 | 1,764 | 25,072 | 84 |
| **Mean** | 21,826 | 4,844 | 93 | 1,320 | 4 |

Variants were considered high-quality with a Bionano confidence score greater than 0.99 (Phred quality score of 20). All ONT variants passed the quality criteria (FILTER=PASS) and exhibited a Phred quality score of ≥20. Variants with an allele frequency of ≤0.01 in the internal Bionano control database or the gnomAD 4.1 SV database were considered rare. Rare: AF≤0.01 in the internal Bionano control database or gnomAD SV 4.1.

In the ONT data, the SV caller Sniffles2 and the CNV caller Spectre identified 414,688 SVs and CNVs across all samples with a mean number of 21,826 per individual. All variant calls by Sniffles2 met the quality cutoff and showed a Phred quality score of at least 20. Across all variants detected with ONT, 92,030 were larger than 500 bp (resolution threshold of OGM). Subsequently, 1,764 variants were in known genes associated with movement disorders. Considering population variant frequency, 25,072 variants showed at least 50% overlap with a gnomAD SV database entry (Overlap fraction product ≥50%) and a rare population allele frequency of ≤1%. Finally, a mean of five variants per sample was both rare in the gnomAD SV database and was found in genes implicated in movement disorders (84 in total over all samples).

### Fractions of structural variant types and length distribution

The fractions of SV types detected by OGM and ONT vary among the different filtered subsets (Figure 2). For both methods, the detected SVs are predominantly deletions and insertions, with a combined fraction of 97.6-99.6% in all subsets. Duplications (0.0-1.3%), inversions (0.0-1.0%), and translocations (0.0-0.4%) are less represented (Supplementary Table 3).

**Figure 2:**
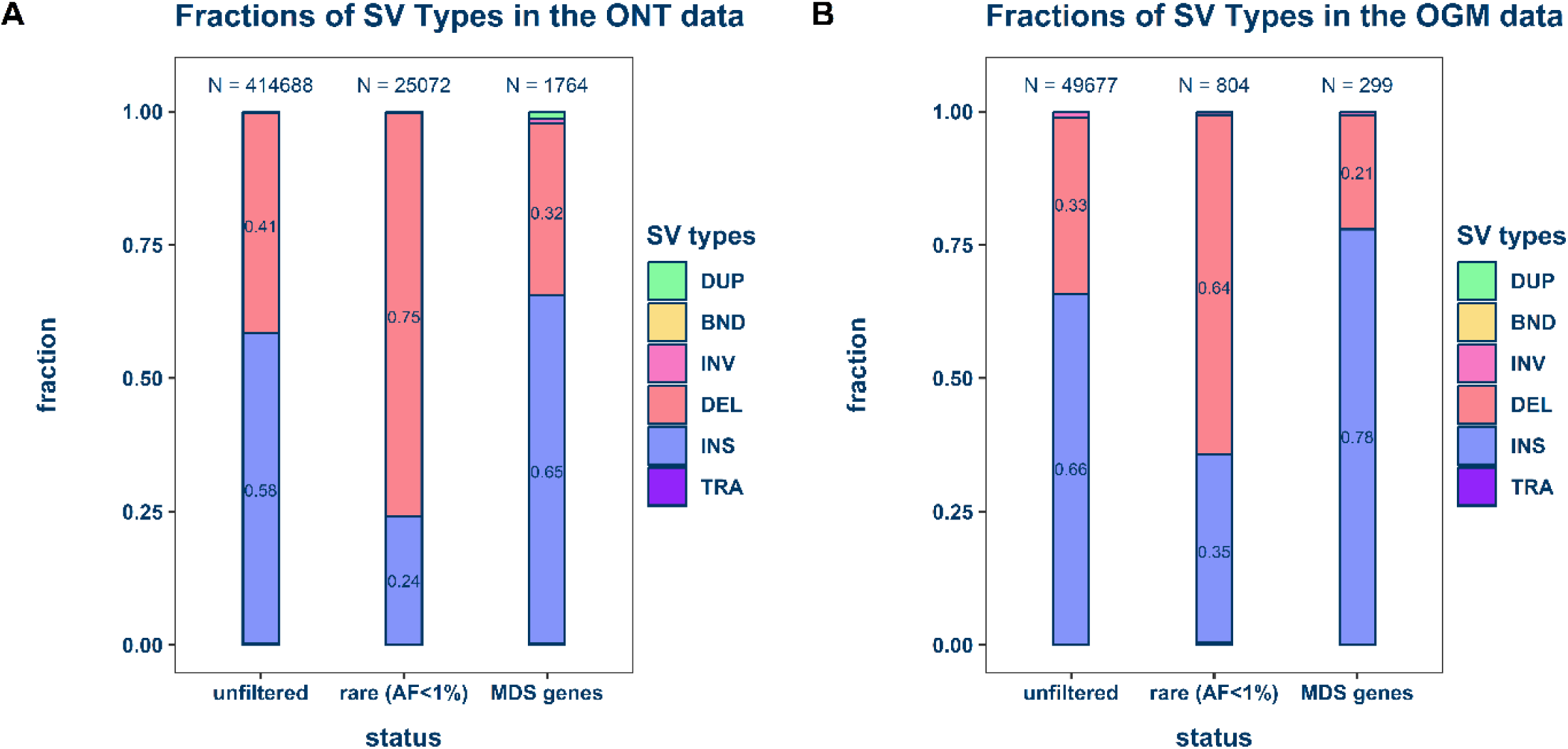
Fraction of SV and CNV types. The stacked bar plots show the fraction of duplications (DUP), break-ends (BND), inversions (INV), deletions (DEL), insertions (INS), and translocations (TRA) in all SVs and CNVs identified by OGM and ONT LRS and in a subset of variants overlapping with known genes associated with movement disorders. **A**. Unfiltered and MDS variants identified in the ONT data. **B**. Unfiltered and MDS variants identified in the OGM data.

The high-confidence variants detected by OGM consist of 32,706 (65.8%) insertions and 16,422 (33.1%) deletions (Figure 2B). In comparison, 284 (35.3%) insertions and 512 (63.7%) deletions are in the rare subset. The variants in movement disorder genes (25) consisted of 233 (78.0%) insertions and 64 (21.4%) deletions. ONT detected 214,972 (58.4%) insertions and 170,953 (41.2%) deletions (Figure 2A). In comparison, the subset of rare variants consists of 6,062 (24.2%) insertions and 18,921 (75.5%) deletions. The variants in genes implicated in movement disorders comprise 1,151 (65.3%) insertions and 570 (32.3%) deletions. The fraction of deletions in the rare subset was overrepresented for both ONT (from 33.1% to 63.7%) and OGM (from 41.1% to 75.5%).

Both methods display a distinct length distribution of all insertions and deletions exceeding 500 bp (Figure 3). In the distribution of the ONT variants (Figure 3A), the variants called by Sniffles2 are, except for a small number of very large SVs, mostly confined to smaller sizes (≤50 kb). Spectre called larger CNVs starting at approximately 80 kb. Only six SVs are detected in the length bracket of approximately 50-80 kb for both insertions and deletions. This visible detection gap is not present in the distribution of the OGM variants; 384 SVs are detected, including the six SVs detected by ONT (Figure 3B). OGM detected a mean deletion length of 29,357 bp, approximately five times larger than that of the ONT variants (6,036 bp). The standard deviation was slightly higher for the OGM variants (±229,773 bp compared to ±177,950 bp), and the median deletion size of the OGM calls was about twice that of the ONT variants (2,994 bp compared to 1,389 bp). The mean insertion length of the ONT variants was slightly larger (6,542 bp compared to 5,807 bp), and the median was approximately half the length (1,093 bp compared to 2,367 bp). The standard deviation of the ONT insertion calls was much higher (±650,874 bp compared to ±18,948 bp). Across all variant types, OGM detected fewer SVs (49,677) with a slightly larger mean size of 25 kb (SD=209 kb) compared to ONT (92,030, mean=17 kb, SD=1.1 Mb).

**Figure 3:**
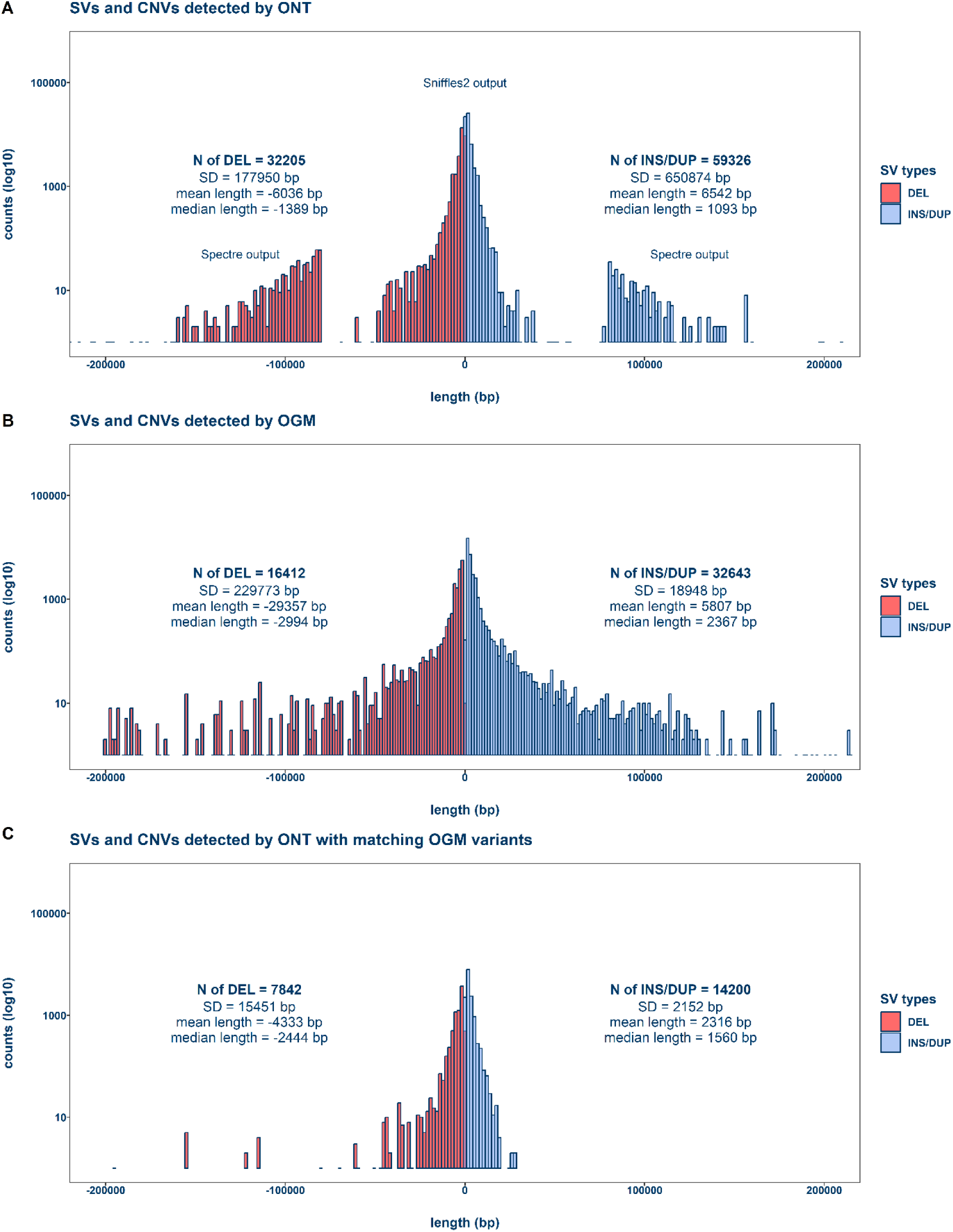
Length distribution of SVs and CNVs. The mirrored histograms show the length distribution of deletions and insertions/duplications ≥500 bp called by ONT and OGM. The variant length in base pairs (bp) is plotted against the logarithmic counts of the variants. **A**: SVs and CNVs identified by Sniffles2 and Spectre in the ONT data. **B**: SVs and CNVs identified by Bionano Solve in the OGM data. **C**: Variants called by ONT with overlapping calls by OGM. DEL: deletion, DUP: duplication, INS: insertion.

Wilcoxon rank-sum tests on the length distribution of both methods (Figure 4) revealed that insertions and deletions detected by OGM were significantly larger than those detected by ONT (p-value <2.2×10^-16^). An empirical cumulative distribution function (ECDF) of the distributions further illustrates this, as the OGM function is shifted towards longer SV lengths for all insertions and deletions (Supplementary Figure 2). Most deletions greater than 50 kb detected by ONT are CNVs of a length of approximately 80-150 kb, with a smaller number of very long (up to 22 Mb) deletions (Figure 4A, D). Spectre and Sniffles2 identified 253 and 31 duplications over 50 kb, respectively, in the size range of up to ∼1 Mb, similar to the OGM insertions. Five duplications were large outliers, measuring up to 149 Mb (Figure 4B). However, Sniffles2 called no clear insertions >50 kb in the ONT data. A characteristic peak of LINE1 mobile elements at approximately 6 kb (43, 44) is visible for both deletions and insertions.

**Figure 4:**
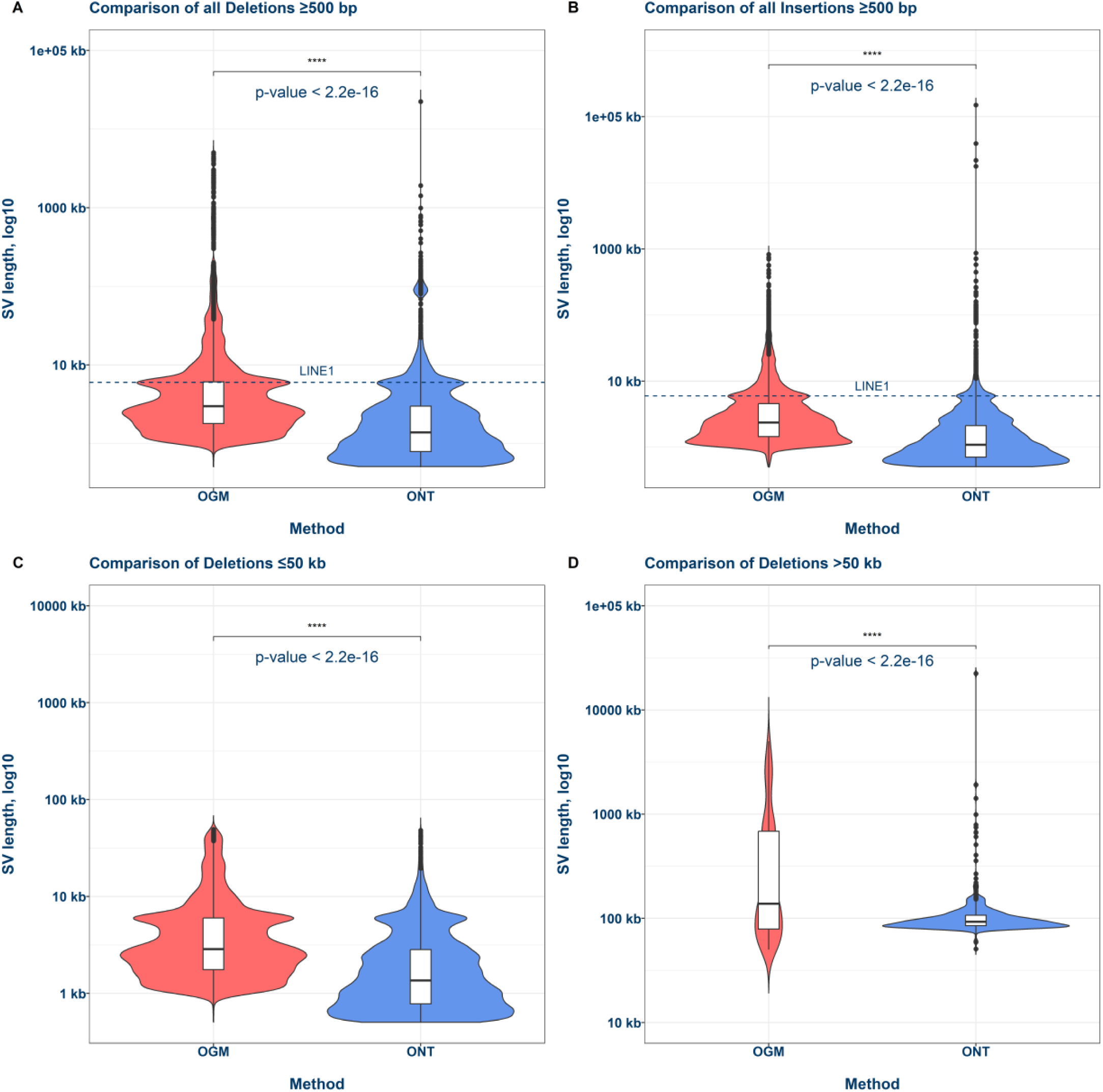
Comparison of SV and CNV lengths. Violin plots and box plots (median) illustrate the length distribution of SVs and CNVs detected by OGM and ONT. p-value: **A** Wilcoxon signed-rank test was performed using the SV length vectors of both methods. A: Comparison of all deletions over the size of 500 bp. **B**: Comparison of all insertions over the size of 500 bp, no insertions over 50 kb were detected by ONT. **C**: Comparison of all deletions ≤50kb. **D**: Comparison of all deletions >50 kb. The dashed line indicates a peak of LINE1 mobile elements near ±6 kb.

### Concordance between OGM and ONT

Using a fixed interval of 6 kb for small (≤50 kb) and 15 kb for intermediate/large (>50 kb) deletions and insertions, the concordance of OGM and ONT is low (Figure 5). About twice as many small deletions (ONT: 31,607; OGM: 15,762) and insertions (ONT: 59,042; OGM: 32,147) were called by ONT compared to OGM, with 7,820 deletions (24.7%) and 14,200 insertions (24.1%) having a matching entry in the OGM variants (Figure 5A, B). In the context of large variants (>50 kb), a similar number of deletions was called by OGM and ONT (ONT: 598; OGM: 650), while OGM called about twice as many insertions (ONT: 284; OGM: 496). Only 22 large deletions (3.7%) detected by ONT had a matching entry in the OGM data, while no large insertions showed overlap between both methods (Figure 5C, D). Out of the ONT variants, 2,416 deletions (7.5%) and 5,256 insertions (8.9%) were identified as rare (occurring only once or less) by needLR in the 1KGP-ONT database. Regarding the variants detected by both methods, 243 deletions (3.1%) and 1,153 insertions (8.1%) were considered rare.

**Figure 5:**
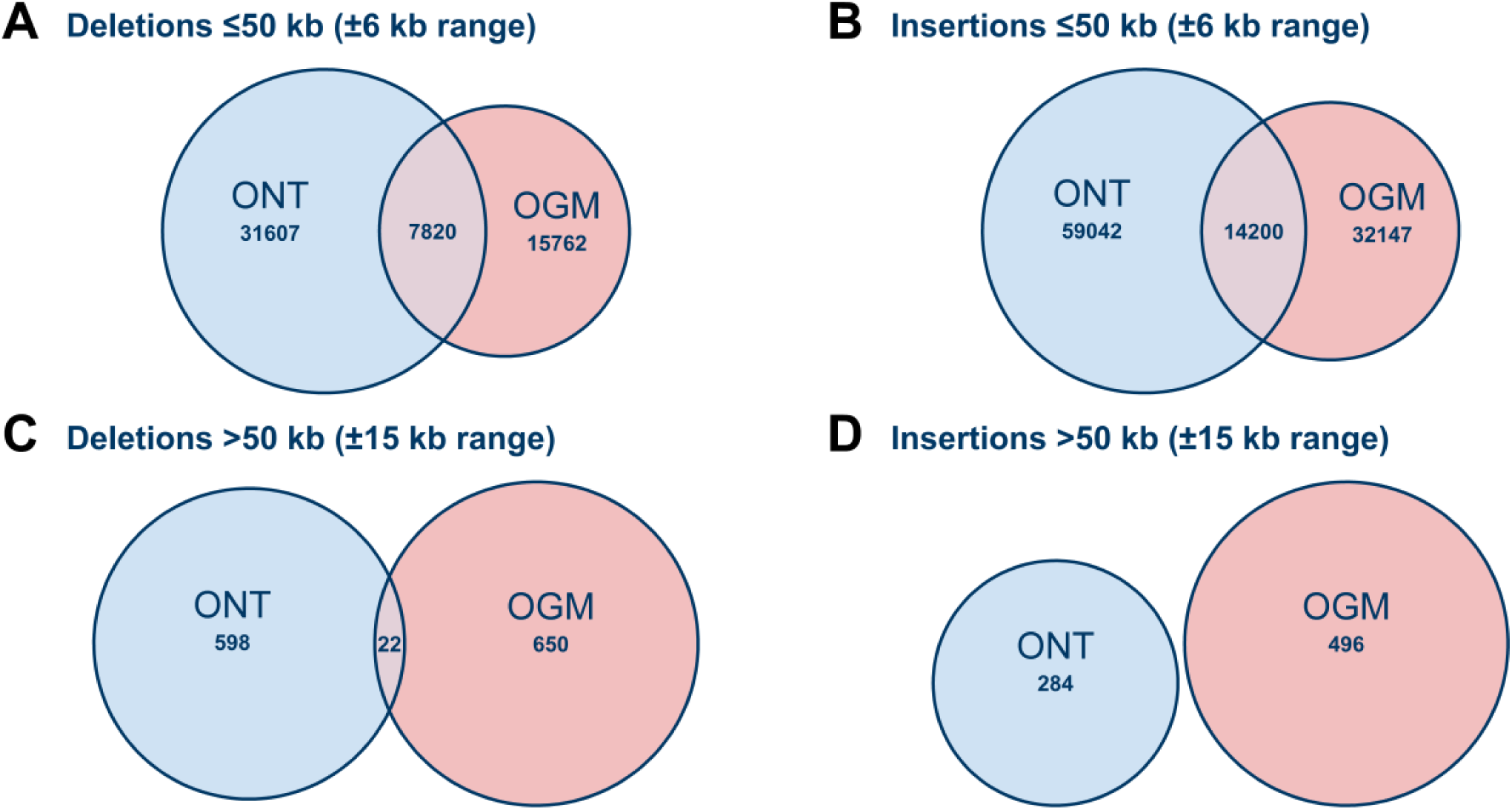
Concordance of variants between ONT and OGM. The Venn diagrams show the approximate overlap between small (>500 bp and ≤50 kb) and large (≥50 kb) deletions (**A**, **C**) and insertions (**B**, **D**) identified by ONT and OGM. Due to the difference in methodology, the SVs in the overlap fraction are not exact matches. A fixed interval (±6 kb and 15 kb) regarding the start and end of the variants was applied based on the median 95% confidence interval of the OGM calls (Supplementary Table 2).

The variants detected with both methods (Figure 5C) are predominantly small, with a mean length of 4,333 bp for the deletions and 2,316 bp for the insertions (SD=15,451 bp and SD=2,152 bp, respectively). However, the 22 large deletions found by both methods (Figure 3C) are visible as discrete bins in the histogram.

### OGM detects large variants in movement disorder genes

Both methods detected a similar number of deletions (ONT: 77, OGM: 64) and insertions (ONT: 307, OGM: 232) in known genes associated with movement disorders (Supplementary Figure 3). Out of the ONT calls, 39 deletions (50.6%) and 56 insertions (19.7%) have a matching entry in the OGM variants. There are differences in the length distribution of the variants in movement disorder genes (Supplementary Figure 4). ONT detected a mean deletion length of 16,737 bp (SD: ±36,130 bp) and a mean insertion length of 674,405 bp (SD: ±8,862,507 bp) (Supplementary Figure 4A). OGM detected a mean deletion length of 242,543 bp (SD: ±456,468 bp) and a mean insertion length of 3,674 bp (SD: ±7,936 bp) (Supplementary Figure 4B). However, the median deletion length (4,993 bp) and insertion length (2,192 bp) of the OGM variants are twice as high as those of the ONT variants (2,375 bp and 1,121 bp, respectively). The number of variants matching between the two methods are small (Supplementary Figure 4C), with a mean deletion length of 2,637 bp (SD: ±2,031 bp) and a mean insertion length of 1,746 bp (SD: ±504 bp).

### ONT refines variant positions in movement disorder genes

Specific variants were evaluated in more detail to highlight and compare the aptitudes of both methods. Three small heterozygous variants in the movement disorder genes *ATXN2*, *SUCLA2*, and *PNKD* showed a rare allele frequency in the internal Bionano control database (Figure 6). OGM detected the heterozygous 1.4 kb deletion in *ATXN2* (Figure 6A) at the position chr12:111,591,863–111,601,607 with an uncertainty (95% CI) of ±4,157 bp. This region spans exon 1 of the *ATXN2* gene. OGM detected the 2.6 kb deletion in *PNKD* (Figure 6B) at the position chr2:218,327,991–218,334,781 with an uncertainty (95% CI) of ±2,074 bp. This region only spans intronic sequence. The heterozygous 1.7 kb deletion in *SUCLA2* (Figure 6C) was detected by OGM at the position chr13:47,967,510–47,980,047 with a 95% confidence interval (CI) of ±5,414 bp. This region overlaps with exons 5 and 6 of *SUCLA2*. All three variants were also identified by ONT (Supplementary Figures 5, 6, and 7) with high precision (95% CI of ±0 bp) and were found to be rare in the 450 current samples of the 1KGP-ONT consortium; only the *ATXN2* SV was found in one sample of the database (HG01372). Yet, all three variants were specified as affecting only intronic sequences.

**Figure 6:**
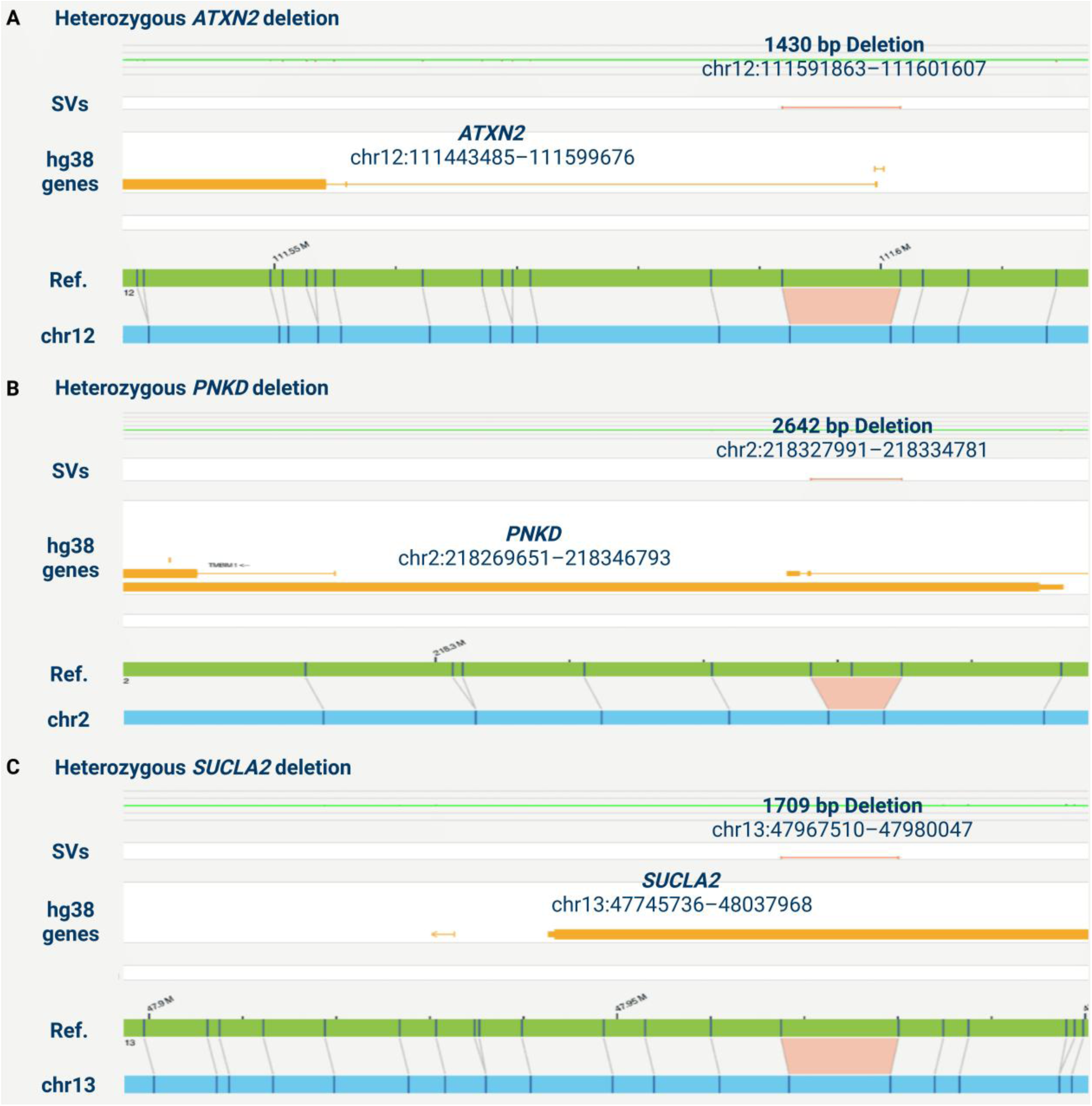
Bionano Access snapshots of three deletions affecting MDS Taskforce genes. The figure shows three small heterozygous deletions in known genes associated with movement disorders identified by both OGM and ONT. All of them lie in intronic regions. **A**: A 1.4 kb deletion in the individual IND-2 affecting the *ATXN2* gene. **B**: A 2.6 kb deletion in IND-17 affecting *PNKD*. **C**: A 1.7 kb deletion in IND-12 affecting *SUCLA2*. Created in BioRender. Fienemann, A. (2025).

Both OGM (Figure 7A) and ONT (Figure 7B) identified a large heterozygous deletion of 195 kb in an intergenic region near the gene *ITPR1*. OGM detects the variant with an uncertainty (95% CI) of ±1,972 bp, and an apparent reduction in the copy number is observed in the same area. The variant showed an allele frequency of <1% in the internal Bionano SV database, gnomAD SVs v4.1.0, and the 450 current samples of the 1KGP-ONT consortium. A PHRED-scaled CADD-SV score of 21.1 was obtained for the variant, indicating that it is in the top 1% of variants in the gnomAD-SV score distribution.

**Figure 7:**
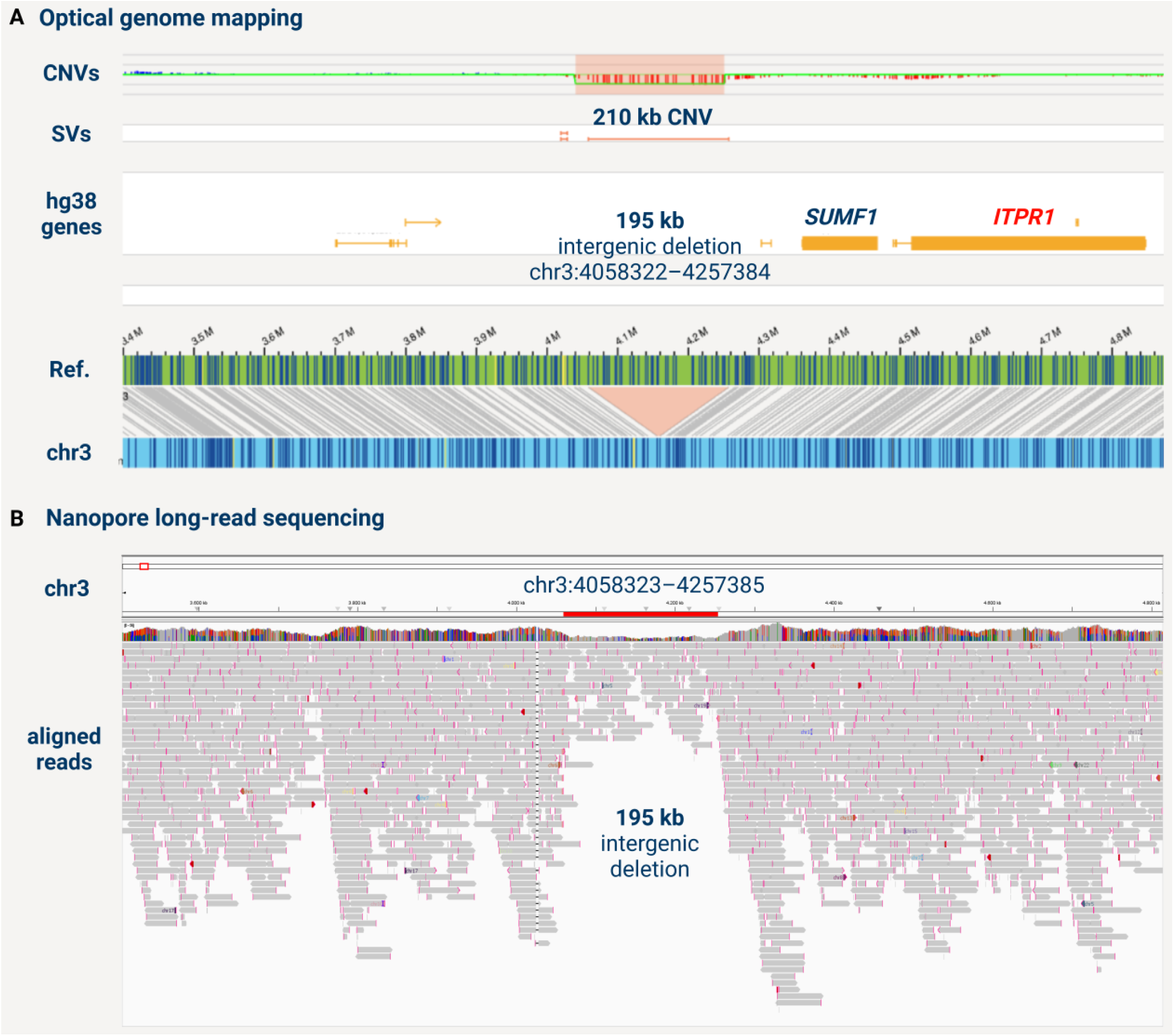
195 kb intergenic deletion. The figure shows a heterozygous 195 kb intergenic deletion near *ITPR1*, detected by both OGM and ONT. **A**: A Bionano Access snapshot of the variant, showing a reduced copy number in the same region. **B**: A snapshot from the Integrative Genomics Viewer (IGV) displaying ONT reads mapped to the GRCh38 reference genome. Created in BioRender. Fienemann, A. (2025).

### Case study: Large PRKN inversion detection by OGM

Finally, a sample with a previously reported 7 Mb inversion affecting *PRKN*, detected using ONT in two monozygotic twins with young-onset dystonia-parkinsonism (16), was analyzed using OGM to test the method’s capabilities further. An inverted intrachromosomal translocation affecting exon 12 of *PRKN* and an exon 3 deletion in trans were detected (Figure 8). The breakpoints of the translocation were detected at the GRCh38 positions 5’: chr6:161,351,957 and 3’: chr6:168,777,862. The *PRKN* deletion was detected at the position chr6:162,205,394-162,278,131.

**Figure 8:**
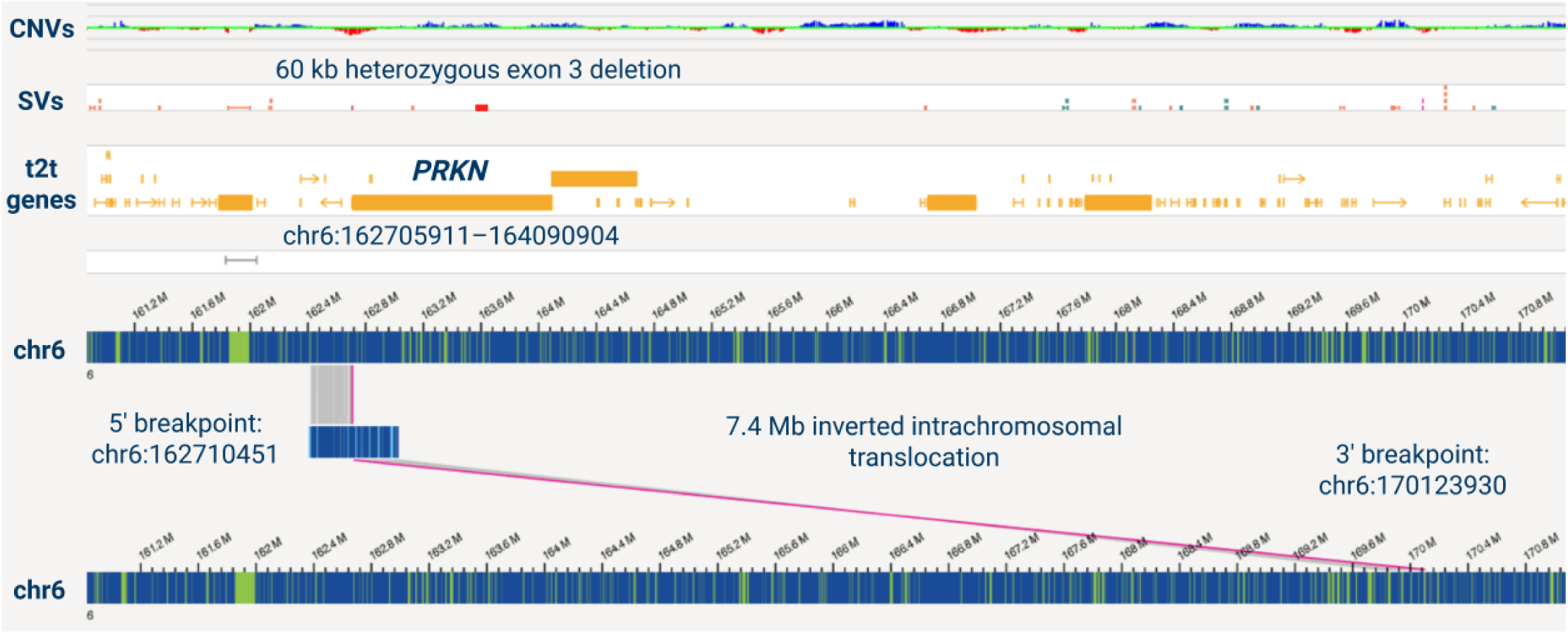
Bionano Access snapshot of a 7 Mb inversion affecting *PRKN*. The figure shows a 7 Mb inversion originally discovered in monozygotic twins with young onset dystonia-parkinsonism (16). Both the large inversion and the exon 3 *PRKN* deletion in trans are detected by optical genome mapping. Due to the size of the variant, it is called as an inverted intrachromosomal translocation. Created in BioRender. Fienemann, A. (2025).

## Discussion

In this study, we compared the SV detection capabilities of ONT and OGM in the whole genomes of *N*=19 idiopathic PD patients, evaluating genome-wide concordance and complementarity, as well as their ability to detect potentially pathogenic variants in known movement disorder-associated genes. Both methods identified a total number of SVs per genome within the previously reported range of ∼20,000 SVs and CNVs for ONT (34, 45) and ∼5,000 for OGM (17, 46), which primarily consisted of insertions and deletions. Insertions accounted for the largest percentage of all variants detected by OGM and ONT (approximately 60%). However, their fraction dropped by half in the filtered subset of rare variants, especially for the ONT variants with a gnomAD SV allele frequency ≤1%. As the gnomAD database is based on short-read sequencing, this could indicate its limited capacity to detect insertions compared to deletions. A recently published effort to sequence 1,019 population-diverse human genomes using ONT revealed that 50.9% of identified insertions were absent in gnomAD SV (47). In contrast, only 14.5% of deletions were not previously represented in the short-read-based database. Sequencing-based approaches generally struggle with large insertions, as they disrupt accurate read alignment (10) and pose unique challenges for SV calling algorithms, e.g., due to junctional homologies at the breakpoints (48). LRS has improved SV detection accuracy compared to short-read sequencing, particularly in the detection of insertions (6, 13), highlighting the utility of technology-matched SV catalogs, such as the 1KGP-ONT Consortium. While these hurdles also apply to OGM, the high coverage (>150 X) and long mean molecule length (>200 kb) of the measured molecules even enabled the detection of very large insertions, as demonstrated by the comparison of SV and CNV lengths between the two methods.

For all deletions and insertions, the OGM-detected variants were significantly longer than those detected by ONT (p <2.2×10^-16^). On the other hand, ONT detected a considerably larger number of variants. In addition to small SVs and Indels under the resolution limit of OGM, twice the number of deletions and insertions exceeding 500 bp were detected compared to the stringently filtered OGM variants. While a fraction of the variants could be attributed to artifacts due to limited base-calling accuracy, ONT is highly capable of detecting small and intermediate-length SVs with high precision (12, 14, 49). The length distribution of the ONT variants illustrates the benefit of performing read depth-based CNV calling, in addition to SV calling by Sniffles2, which was previously known to be limited in detecting larger variants (>50 kb) (50). However, there are exceptions, such as the heterozygous 195 kb intergenic deletion near the ataxia gene *ITPR1* (51, 52), which OGM detected and ONT validated with single-base pair accuracy (95% CI of ±0 bp). The phred-scaled CADD-SV score of >20 indicates a moderately predicted deleteriousness of the variant, potentially due to the spanning of cis-regulatory elements. A detection gap remains in the length bracket of approximately 50-80 kb for both insertions and deletions detected by ONT, as CNV calling by Spectre is optimized for variants >100 kb. OGM detected 64 times more variants in this range, including the six SVs detected by ONT. Although the widespread adoption of LRS has led to steady improvements in variant callers, only OGM detected a heterozygous 98 kb deletion spanning the *PEX7* gene (Supplementary Figure 8). Using v2.3.3 of Sniffles2 or using Spectre, ONT did not detect the SV (Supplementary Figure 9) as it falls outside the optimal detection range of both. However, using the recent v2.6.2 of Sniffles2, the variant was detected with high precision (95% CI of ±0 bp) (Supplementary Figure 10).

Previous studies compared SVs detected by ONT and OGM in SKBR3 human breast cancer cells (53), the plant genome of Arabidopsis thaliana (54), 234 individually curated variants in *N*=9 parent-child trios (6), and renal cell carcinoma tumor tissue (55). All of these studies reported a generally high degree of overlap between the two methods, as well as some discrepancies in SV detection due to differences in methodology and aptitude for different variant types and lengths, suggesting a synergy between the two approaches. However, to our knowledge, no previous study has compared the SV detection of ONT and OGM genome-wide in non-neoplastic human-derived samples. In our study, the concordance between OGM and ONT was low, especially for large variants (>50 kb), with ∼3.7-24.7% of ONT variants matching those in the OGM dataset. There were also no differences in the percentage of rare 1KGP variants between the unique ONT SVs and the concordant SVs, indicating that neither technique showed a bias towards common variants. In the most recent comparison of both methods in renal cell carcinoma, the highest overlap was reported for the 1–5 kb length bracket, with 1487 (81.7%) SVs being concordant (55). ONT and OGM detected 1,821 and 1,679 SVs, respectively. For SVs ≥10 kb, OGM detected 232 SVs, compared to 48 detected by ONT, with 40 (∼83.3%) being concordant (55). However, only SVs called by Sniffles2 and no CNVs identified by Spectre or other methods were compared. In the comparison in our study, CNVs were included, resulting in a lower concordance between OGM and ONT due to the reduced breakpoint accuracy and higher false-positive rate of the read depth approach, as 823 out of 986 variants exceeding 50 kb were CNVs called by Spectre. Different versions of base-calling models and variant detection tools for the ONT data also contribute to differences between our study and previous comparisons, e.g., Guppy base-calling instead of Dorado (Pei et al., 2024). Still, a limitation of the here performed comparison is that while a fixed interval regarding the breakpoint positions provides a high-level overview, not all nuances are captured, e.g., subtle differences of smaller SVs or complex loci. This also reveals a limitation of OGM, being the breakpoint accuracy, which showed high variance depending on the local label density, with a median 95% CI of 15 kb (SD = 41 kb) for variants greater than 50 kb.

Catalogs of challenging and medically relevant genes, such as the ACMG (56) or other reported gene lists, e.g., the Genome in a Bottle challenging medically relevant genes benchmark set of 217 SVs (57, 58), were helpful in the past to highlight the power of long-reads to overcome repetitive gene regions and pseudogene homology (59). Here, we studied the ability of OGM and ONT to identify medically relevant variants in known genes associated with movement disorders presented by the MDS Taskforce (25). Both methods detected the same three rare and heterozygous deletions in the genes *ATXN2*, *SUCLA2*, and *PNKD*. *ATXN2* is implicated in spinocerebellar ataxia (60) but can also present with predominant Parkinsonism (61), while *PNKD* can paroxysmal non-kinesigenic dyskinesia (62). Both show autosomal dominate inheritance. *SUCLA2* can cause complex dystonia (63, 64) with autosomal recessive inheritance. Although OGM detected all three variants with high confidence, the uncertainty in the exact breakpoint positions prevented practical interpretation of the variants, as it was unclear if the coding sequence was involved or not. ONT finally confirmed all three to be intronic, emphasizing the advantage of single-basepair resolution for interpreting, especially smaller SVs. In the Japanese PD sample, OGM detected both the previously identified 7.4 Mb inverted intrachromosomal translocation and 60 kb deletion in trans (16) affecting the early-onset PD gene *PRKN* (65). This aligns with previous observations that the Bionano Solve software represents inversions exceeding 5 Mb due to their size as intrachromosomal translocations (66). OGM was able to phase the known inversion and deletion in trans and validate the compound heterozygous disruption of *PRKN* in a straightforward manner, confirming that OGM excels in detecting of large and complex SVs.

## Conclusions

The study of SVs has great potential to unravel the missing genetic factors and monogenic causes in the field of PD and other neurodegenerative disorders. Technologies like LRS and OGM provide us with the means to fill the gaps left by established methods, such as NGS (67). This is highlighted in a recent study where six individuals with unsolved SVs were analyzed using both ONT and OGM to resolve breakpoint junctions at the single-basepair level and allow for effective variant phasing to unravel complex rearrangements (22). As OGM has a high aptitude for detecting especially large and complex variants, and ONT excels in characterizing smaller to intermediate-sized variants, their combined application offers a complementary approach to enable broad detection and robust validation of structural variation.

## Declarations

### Ethics approval and consent to participate

Written informed consent was obtained from all individuals and approved by the Ethics Committee at the University of Lübeck, Lübeck, Germany, and the University of British Columbia, Vancouver, Canada.

### Consent for publication

Not applicable.

### Availability of data and materials

The datasets used and/or analyzed during the current study are available from the corresponding author on reasonable request.

### Authors’ contributions

AF contributed to the conceptualization of the study, data curation, formal analysis, methodology, investigation, software, visualization, writing of the original draft, and final approval of the manuscript. TL was involved in conceptualization, data curation, methodology, software, supervision, and final approval of the manuscript. SS participated in the methodology, investigation, and final approval of the manuscript. CG and MM contributed to methodology, data curation, software, and final approval of the manuscript. HB and KL contributed to funding acquisition, and final approval of the manuscript. JAG and DEM contributed to the provision of software resources, and final approval of the manuscript. KD, MF, NH, SBS, FH, and MJF provided patient samples, clinical information of patients, and read and approved the final manuscript. KKU participated in the investigation, software, and final approval of the manuscript. CK contributed to the conceptualization, project administration, funding acquisition, provision of resources, and final approval of the manuscript. JT was involved in conceptualization, funding acquisition, project administration, resource provision, supervision, and final approval of the manuscript.

## Acknowledgments

We extend our sincere gratitude to the patients and their families for their participation in this study and their generous donation of biospecimen samples.

## References

1. De Coster W, Van Broeckhoven C. Newest Methods for Detecting Structural Variations. Trends Biotechnol. 2019;37(9):973–82.

2. Collins RL, Talkowski ME. Diversity and consequences of structural variation in the human genome. Nat Rev Genet. 2025;26(7):443–62.

3. Gustavsson EK, Follett J, Trinh J, Barodia SK, Real R, Liu Z, et al. RAB32 Ser71Arg in autosomal dominant Parkinson’s disease: linkage, association, and functional analyses. Lancet Neurol. 2024;23(6):603–14.

4. Trinh J, Lohmann K, Baumann H, Balck A, Borsche M, Bruggemann N, et al. Utility and implications of exome sequencing in early-onset Parkinson’s disease. Mov Disord. 2019;34(1):133–7.

5. Petersen BS, Fredrich B, Hoeppner MP, Ellinghaus D, Franke A. Opportunities and challenges of whole-genome and -exome sequencing. BMC Genet. 2017;18(1):14.

6. Pei Y, Tanguy M, Giess A, Dixit A, Wilson LC, Gibbons RJ, et al. A Comparison of Structural Variant Calling from Short-Read and Nanopore-Based Whole-Genome Sequencing Using Optical Genome Mapping as a Benchmark. Genes (Basel). 2024;15(7).

7. Kernohan KD, Boycott KM. The expanding diagnostic toolbox for rare genetic diseases. Nat Rev Genet. 2024;25(6):401–15.

8. Eisfeldt J, Ek M, Nordenskjold M, Lindstrand A. Toward clinical long-read genome sequencing for rare diseases. Nat Genet. 2025;57(6):1334–43.

9. Espinosa E, Bautista R, Larrosa R, Plata O. Advancements in long-read genome sequencing technologies and algorithms. Genomics. 2024;116(3):110842.

10. Mahmoud M, Gobet N, Cruz-Davalos DI, Mounier N, Dessimoz C, Sedlazeck FJ. Structural variant calling: the long and the short of it. Genome Biol. 2019;20(1):246.

11. Alvarez Jerez P, Wild Crea P, Ramos DM, Gustavsson EK, Radefeldt M, Damianov A, et al. African ancestry neurodegeneration risk variant disrupts an intronic branchpoint in GBA1. Nat Struct Mol Biol. 2024;31(12):1955–63.

12. Cogan G, Daida K, Billingsley KJ, Tesson C, Forlani S, Jornea L, et al. Long-Read Sequencing Unravels the Complexity of Structural Variants in PRKN in Two Individuals with Early-Onset Parkinson’s Disease. Mov Disord. 2024;39(9):1647–8.

13. Billingsley KJ, Ding J, Jerez PA, Illarionova A, Levine K, Grenn FP, et al. Genome-Wide Analysis of Structural Variants in Parkinson Disease. Ann Neurol. 2023;93(5):1012–22.

14. Daida K, Yoshino H, Malik L, Baker B, Ishiguro M, Genner R, et al. The Utility of Long-Read Sequencing in Diagnosing Early Onset Parkinson’s Disease. Ann Neurol. 2025;97(4):753–65.

15. Miano-Burkhardt A, Alvarez Jerez P, Daida K, Bandres Ciga S, Billingsley KJ. The Role of Structural Variants in the Genetic Architecture of Parkinson’s Disease. Int J Mol Sci. 2024;25(9).

16. Daida K, Funayama M, Billingsley KJ, Malik L, Miano-Burkhardt A, Leonard HL, et al. Long-Read Sequencing Resolves a Complex Structural Variant in PRKN Parkinson’s Disease. Mov Disord. 2023;38(12):2249–57.

17. Mantere T, Neveling K, Pebrel-Richard C, Benoist M, van der Zande G, Kater-Baats E, et al. Optical genome mapping enables constitutional chromosomal aberration detection. Am J Hum Genet. 2021;108(8):1409–22.

18. Alkan C, Coe BP, Eichler EE. Genome structural variation discovery and genotyping. Nat Rev Genet. 2011;12(5):363–76.

19. Facchini S, Dominik N, Manini A, Efthymiou S, Curro R, Rugginini B, et al. Optical Genome Mapping Enables Detection and Accurate Sizing of RFC1 Repeat Expansions. Biomolecules. 2023;13(10).

20. Ghorbani F, de Boer-Bergsma J, Verschuuren-Bemelmans CC, Pennings M, de Boer EN, Kremer B, et al. Prevalence of intronic repeat expansions in RFC1 in Dutch patients with CANVAS and adult-onset ataxia. J Neurol. 2022;269(11):6086–93.

21. Dremsek P, Schachner A, Reischer T, Krampl-Bettelheim E, Bettelheim D, Vrabel S, et al. Retrospective study on the utility of optical genome mapping as a follow-up method in genetic diagnostics. J Med Genet. 2025;62(2):89–96.

22. De Clercq G, Vantomme L, Dewaele B, Callewaert B, Vanakker O, Janssens S, et al. Full characterization of unresolved structural variation through long-read sequencing and optical genome mapping. Sci Rep. 2024;14(1):29142.

23. Sedlazeck FJ, Lee H, Darby CA, Schatz MC. Piercing the dark matter: bioinformatics of long-range sequencing and mapping. Nat Rev Genet. 2018;19(6):329–46.

24. Trinh J, Schaake S, Gabbert C, Luth T, Cowley SA, Fienemann A, et al. Optical genome mapping of structural variants in Parkinson’s disease-related induced pluripotent stem cells. BMC Genomics. 2024;25(1):980.

25. Lange LM, Gonzalez-Latapi P, Rajalingam R, Tijssen MAJ, Ebrahimi-Fakhari D, Gabbert C, et al. Nomenclature of Genetic Movement Disorders: Recommendations of the International Parkinson and Movement Disorder Society Task Force - An Update. Mov Disord. 2022;37(5):905–35.

26. Wenghöfer A, Paquette K, Malik L, Baker B, Kouam C, Billingsley KJ. Processing frozen archival human DNA samples for large-scale SQK-LSK114 Oxford Nanopore long-read DNA sequencingSOP v1 v1. protocolsio. 2024.

27. Li H, Handsaker B, Wysoker A, Fennell T, Ruan J, Homer N, et al. The Sequence Alignment/Map format and SAMtools. Bioinformatics. 2009;25(16):2078–9.

28. Li H. Minimap2: pairwise alignment for nucleotide sequences. Bioinformatics. 2018;34(18):3094–100.

29. Smolka M, Paulin LF, Grochowski CM, Horner DW, Mahmoud M, Behera S, et al. Detection of mosaic and population-level structural variants with Sniffles2. Nat Biotechnol. 2024;42(10):1571–80.

30. Sedlazeck FJ, Rescheneder P, Smolka M, Fang H, Nattestad M, von Haeseler A, et al. Accurate detection of complex structural variations using single-molecule sequencing. Nat Methods. 2018;15(6):461–8.

31. Sedlazeck FJ, Sanio P, Paulin LF. Spectre - Long read CNV caller: GitHub; 2024 [Available from: github.com/fritzsedlazeck/Spectre.

32. Geoffroy V, Herenger Y, Kress A, Stoetzel C, Piton A, Dollfus H, et al. AnnotSV: an integrated tool for structural variations annotation. Bioinformatics. 2018;34(20):3572–4.

33. Nicholas TJ, Cormier MJ, Quinlan AR. Annotation of structural variants with reported allele frequencies and related metrics from multiple datasets using SVAFotate. BMC Bioinformatics. 2022;23(1):490.

34. Gustafson JA, Gibson SB, Damaraju N, Zalusky MPG, Hoekzema K, Twesigomwe D, et al. High-coverage nanopore sequencing of samples from the 1000 Genomes Project to build a comprehensive catalog of human genetic variation. Genome Res. 2024;34(11):2061–73.

35. Kleinert P, Kircher M. A framework to score the effects of structural variants in health and disease. Genome Res. 2022;32(4):766–77.

36. R Core Team. R: A Language and Environment for Statistical Computing. R Foundation for Statistical Computing, Vienna, Austria. 2025.

37. Barrett T, Dowle M, Srinivasan A, Gorecki J, Chirico M, Hocking T. data.table: Extension of ‘data.fram’. R package version 1154. 2024.

38. Wickham H, Averick M, Bryan J, Chang W, McGowan L, François R, et al. Welcome to the Tidyverse. Journal of Open Source Software. 2019;4(43):1686.

39. Wickham H, François R, Henry L, Müller K, Vaughan D. dplyr: A Grammar of Data Manipulation. R package version 114. 2023.

40. Wickham H. ggplot2: Elegant Graphics for Data Analysis. Springer-Verlag New York. 2016.

41. Richards S, Aziz N, Bale S, Bick D, Das S, Gastier-Foster J, et al. Standards and guidelines for the interpretation of sequence variants: a joint consensus recommendation of the American College of Medical Genetics and Genomics and the Association for Molecular Pathology. Genet Med. 2015;17(5):405–24.

42. Lam ET, Hastie A, Lin C, Ehrlich D, Das SK, Austin MD, et al. Genome mapping on nanochannel arrays for structural variation analysis and sequence assembly. Nat Biotechnol. 2012;30(8):771–6.

43. Zook JM, Hansen NF, Olson ND, Chapman L, Mullikin JC, Xiao C, et al. A robust benchmark for detection of germline large deletions and insertions. Nat Biotechnol. 2020;38(11):1347–55.

44. Collins RL, Brand H, Karczewski KJ, Zhao X, Alfoldi J, Francioli LC, et al. A structural variation reference for medical and population genetics. Nature. 2020;581(7809):444-51.

45. Beyter D, Ingimundardottir H, Oddsson A, Eggertsson HP, Bjornsson E, Jonsson H, et al. Long-read sequencing of 3,622 Icelanders provides insight into the role of structural variants in human diseases and other traits. Nat Genet. 2021;53(6):779–86.

46. Iqbal MA, Broeckel U, Levy B, Skinner S, Sahajpal NS, Rodriguez V, et al. Multisite Assessment of Optical Genome Mapping for Analysis of Structural Variants in Constitutional Postnatal Cases. J Mol Diagn. 2023;25(3):175–88.

47. Schloissnig S, Pani S, Ebler J, Hain C, Tsapalou V, Soylev A, et al. Structural variation in 1,019 diverse humans based on long-read sequencing. Nature. 2025.

48. Delage WJ, Thevenon J, Lemaitre C. Towards a better understanding of the low recall of insertion variants with short-read based variant callers. BMC Genomics. 2020;21(1):762.

49. De Coster W, De Rijk P, De Roeck A, De Pooter T, D’Hert S, Strazisar M, et al. Structural variants identified by Oxford Nanopore PromethION sequencing of the human genome. Genome Res. 2019;29(7):1178–87.

50. Cuenca-Guardiola J, de la Morena-Barrio B, Garcia JL, Sanchis-Juan A, Corral J, Fernandez-Breis JT. Improvement of large copy number variant detection by whole genome nanopore sequencing. J Adv Res. 2023;50:145–58.

51. Iwaki A, Kawano Y, Miura S, Shibata H, Matsuse D, Li W, et al. Heterozygous deletion of ITPR1, but not SUMF1, in spinocerebellar ataxia type 16. J Med Genet. 2008;45(1):32–5.

52. van de Leemput J, Chandran J, Knight MA, Holtzclaw LA, Scholz S, Cookson MR, et al. Deletion at ITPR1 underlies ataxia in mice and spinocerebellar ataxia 15 in humans. PLoS Genet. 2007;3(6):e108.

53. Savara J, Novosad T, Gajdos P, Kriegova E. Comparison of structural variants detected by optical mapping with long-read next-generation sequencing. Bioinformatics. 2021;37(20):3398–404.

54. Canaguier A, Guilbaud R, Denis E, Magdelenat G, Belser C, Istace B, et al. Oxford Nanopore and Bionano Genomics technologies evaluation for plant structural variation detection. BMC Genomics. 2022;23(1):317.

55. Margalit S, Tulpova Z, Detinis Zur T, Michaeli Y, Deek J, Nifker G, et al. Long-read structural and epigenetic profiling of a kidney tumor-matched sample with nanopore sequencing and optical genome mapping. NAR Genom Bioinform. 2025;7(1):lqae190.

56. Miller DT, Lee K, Chung WK, Gordon AS, Herman GE, Klein TE, et al. ACMG SF v3.0 list for reporting of secondary findings in clinical exome and genome sequencing: a policy statement of the American College of Medical Genetics and Genomics (ACMG). Genet Med. 2021;23(8):1381–90.

57. Wagner J, Olson ND, Harris L, McDaniel J, Cheng H, Fungtammasan A, et al. Curated variation benchmarks for challenging medically relevant autosomal genes. Nat Biotechnol. 2022;40(5):672–80.

58. Mandelker D, Schmidt RJ, Ankala A, McDonald Gibson K, Bowser M, Sharma H, et al. Navigating highly homologous genes in a molecular diagnostic setting: a resource for clinical next-generation sequencing. Genet Med. 2016;18(12):1282–9.

59. Mahmoud M, Huang Y, Garimella K, Audano PA, Wan W, Prasad N, et al. Utility of long-read sequencing for All of Us. Nat Commun. 2024;15(1):837.

60. Pulst SM, Nechiporuk A, Nechiporuk T, Gispert S, Chen XN, Lopes-Cendes I, et al. Moderate expansion of a normally biallelic trinucleotide repeat in spinocerebellar ataxia type 2. Nat Genet. 1996;14(3):269–76.

61. Shan DE, Soong BW, Sun CM, Lee SJ, Liao KK, Liu RS. Spinocerebellar ataxia type 2 presenting as familial levodopa-responsive parkinsonism. Ann Neurol. 2001;50(6):812–5.

62. Rainier S, Thomas D, Tokarz D, Ming L, Bui M, Plein E, et al. Myofibrillogenesis regulator 1 gene mutations cause paroxysmal dystonic choreoathetosis. Arch Neurol. 2004;61(7):1025–9.

63. Carrozzo R, Dionisi-Vici C, Steuerwald U, Lucioli S, Deodato F, Di Giandomenico S, et al. SUCLA2 mutations are associated with mild methylmalonic aciduria, Leigh-like encephalomyopathy, dystonia and deafness. Brain. 2007;130(Pt 3):862–74.

64. Ostergaard E, Hansen FJ, Sorensen N, Duno M, Vissing J, Larsen PL, et al. Mitochondrial encephalomyopathy with elevated methylmalonic acid is caused by SUCLA2 mutations. Brain. 2007;130(Pt 3):853–61.

65. Kitada T, Asakawa S, Hattori N, Matsumine H, Yamamura Y, Minoshima S, et al. Mutations in the parkin gene cause autosomal recessive juvenile parkinsonism. Nature. 1998;392(6676):605-8.

66. Gao H, Xu H, Wang C, Cui L, Huang X, Li W, et al. Optical Genome Mapping for Comprehensive Assessment of Chromosomal Aberrations and Discovery of New Fusion Genes in Pediatric B-Acute Lymphoblastic Leukemia. Cancers (Basel). 2022;15(1).

67. Cogan G, Daida K, Blauwendraat C, Billingsley K, Brice A. Exploration of Neurodegenerative Diseases Using Long-Read Sequencing and Optical Genome Mapping Technologies. Mov Disord. 2025;40(6):996–1008.

