## Supplementary tables and figures for "Complementarity of long-read sequencing and optical genome mapping in Parkinson’s disease"

### Supplementary material

**Supplementary Table 1**. Clinical details and demographics of the research cohort.

| Individuals | Sex | Age (2025) | AAO | FH |
|---|---|---|---|---|
| IND-1 | m | 56-60 | 31-35 | negative |
| IND-2 | m | 71-75 | 31-35 | negative |
| IND-3 | m | 66-70 | 31-35 | negative |
| IND-4 | f | 71-75 | 41-45 | positive |
| IND-5 | m | 56-60 | 36-40 | negative |
| IND-6 | f | 61-65 | 46-50 | *NA* |
| IND-7 | m | 51-55 | 36-40 | *NA* |
| IND-8 | m | 71-75 | 46-50 | *NA* |
| IND-9 | m | 56-60 | 36-40 | *NA* |
| IND-10 | f | 41-45 | 41-45 | *NA* |
| IND-11 | m | 66-70 | 36-40 | positive |
| IND-12 | f | 61-65 | 31-35 | *NA* |
| IND-13 | m | 51-55 | 36-40 | *NA* |
| IND-14 | m | 56-60 | 21-25 | *NA* |
| IND-15 | m | 66-70 | 11-15 | positive |
| IND-16 | m | 56-60 | 16-20 | positive |
| IND-17 | f | 56-60 | 36-40 | positive |
| IND-18 | f | 91-95 | 66-70 | positive |
| IND-19 | m | 91-95 | 71-75 | positive |
| **total** | **male (%)** | **Mean±SD** | **Mean±SD** | **total** |
| *N*=19 | 68,42% | 65±13 | 39±14 | *N*=6 |

AAO: age at onset; FH: family history.

**Supplementary Table 2**. Overview of the 95% Confidence interval statistics of the start/end position for the OGM and ONT variants ≥50 kb and ≤50 kb.

| **Optical genome mapping (OGM)** | | | | | |
|---|---|---|---|---|---|
| Mean 95% CI (≤50 kb) | Median 95% CI (≤50 kb) | SD Median 95% CI (≤50 kb) | Mean 95% CI (≥50 kb) | Median 95% CI (≥50 kb) | SD Median 95% CI (≥50 kb) |
| 7,916 bp | 5,698 bp | ±8040 bp | 26,254 bp | 15,052 bp | ±40,948 bp |
| **Nanopore long-read sequencing (ONT)** | | | | | |
| Mean 95% CI (≤50 kb) | Median 95% CI (≤50 kb) | SD Median 95% CI (≤50 kb) | Mean 95% CI (≥50 kb) | Median 95% CI (≥50 kb) | SD Median 95% CI (≥50 kb) |
| 43 bp | 10 bp | ±80 bp | 12 bp | 12 bp | 2 bp |

CI: Confidence interval.

**Supplementary Table 3**. Fraction of SV types detected by ONT and OGM.

**Optical genome mapping (OGM)**

| Status | Insertion (INS) | Deletion (DEL) | Duplication (DUP) | Inversion (INV) | Break end (BND) | Translocation (TRA) |
|---|---|---|---|---|---|---|
| unfiltered | 32,706 (65.8%) | 16,422 (33.1%) | 37 (0.1%) | 509 (1.0%) | 0 (0%) | 3 (0%) |
| rare (AF<1%) | 284 (35.3%) | 512 (63.7%) | 0 (0%) | 5 (0.6%) | 0 (0%) | 3 (0.4%) |
| MDS genes | 233 (78.0%) | 64 (21.4%) | 0 (0%) | 2 (0.7%) | 0 (0%) | 0 (0%) |

**Nanopore long-read sequencing (ONT)**

| Status | Insertion (INS) | Deletion (DEL) | Duplication (DUP) | Inversion (INV) | Break end (BND) | Translocation (TRA) |
|---|---|---|---|---|---|---|
| unfiltered | 241,972 (58.4%) | 170,953 (41.2%) | 446 (0.1%) | 556 (0.1%) | 0 (0%) | 761 (0.2%) |
| rare (AF<1%) | 6,062 (24.2%) | 18,921 (75.5%) | 81 (0.3%) | 8 (0%) | 0 (0%) | 0 (0%) |
| MDS genes | 1,151 (65.3%) | 570 (32.3%) | 23 (1.3%) | 16 (0.9%) | 0 (0%) | 4 (0.2%) |

Rare, AF<1% in gnomAD SV 4.1.

**Supplementary Table 4: OGM molecule statistics**: Overview of different metrics describing the molecules imaged during OGM of the cohort.

**Bionano optical genome mapping**

| ID | Total molecules | Total length (Mb) | Mean length (kb) | Molecule N50 (kb) | Label density (x/100 kb) | Coverage (X) |
|---|---|---|---|---|---|---|
| L-4035 | 2,136,798 | 598,544 | 280.11 | 288.78 | 16.5 | 193.8 |
| L-3047 | 2,211,967 | 592,207 | 267.73 | 273.85 | 15.7 | 191.8 |
| L-3046 | 2,402,779 | 585,143 | 243.53 | 242.99 | 16.2 | 189.5 |
| L-3050 | 2,287,562 | 511,203 | 223.47 | 214.25 | 15.9 | 165.5 |
| L-5502 | 2,362,733 | 512,296 | 216.82 | 207.34 | 15.8 | 165.9 |
| L-4474 | 2,023,237 | 504,252 | 249.23 | 241.12 | 15.8 | 163.3 |
| L-7657 | 1,911,162 | 473,194 | 247.60 | 246.53 | 16.1 | 153.2 |
| L-8302 | 2,384,948 | 521,934 | 218.85 | 211.23 | 15.9 | 169.0 |
| L-23100 | 1,804,644 | 407,804 | 225.98 | 219.89 | 15.3 | 132.1 |
| L-24764 | 2,050,034 | 591,218 | 288.39 | 298.14 | 16.1 | 191.4 |
| L-3041 | 2,384,948 | 521,934 | 218.85 | 211.23 | 15.9 | 169.0 |
| L-3049 | 4,680,246 | 997,025 | 213.03 | 205.44 | 16.3 | 322.8 |
| L-7759 | 2,132,576 | 506,007 | 237.28 | 235.61 | 15.0 | 163.9 |
| L-5964 | 2,009,951 | 488,949 | 243.26 | 244.49 | 14.8 | 156.9 |
| 3007-10 | 2,422,741 | 586,001 | 241.88 | 242.37 | 16.2 | 189.8 |
| 3007-14 | 2,160,768 | 478,292 | 221.35 | 210.17 | 15.9 | 154.9 |
| 3048-28 | 1,922,930 | 430,528 | 223.89 | 214.78 | 16.3 | 139.4 |
| 3049-10 | 2,173,312 | 577,664 | 265.80 | 270.15 | 16.4 | 187.1 |
| 3049-06 | 2,038,671 | 488,601 | 239.67 | 234.36 | 16.5 | 158.2 |
| **Mean** | 2,289,579 | 545,936 | 240.35 | 237.51 | 15.9 | 176.7 |
| **Median** | 2,160,768 | 512,296 | 239.67 | 235.61 | 15.9 | 165. 9 |
| **SD** | 607,168 | 122,522 | 22.11 | 28.221 | 0.5 | 39.7 |

ID: identifier of the sample e.g., L-number, kb: kilobase, Mb: megabase.

**Supplementary Table 5: ONT read statistics**: Overview of different metrics describing the reads measured during ONT long-read sequencing of the cohort.

| Oxford Nanopore long-read sequencing | | | | | |
|---|---|---|---|---|---|
| ID | Total reads | Mean Q-score | Mean length (bp) | Read N50 (bp) | Coverage (X) |
| L-4035 | 11,243,896 | 15.1 | 6,524 | 9,223 | 23.7 |
| L-3047 | 10,400,070 | 15.0 | 10,446 | 16,463 | 35.3 |
| L-3046 | 9,265,455 | 15.1 | 7,852 | 11,885 | 23.6 |
| L-3050 | 13,790,542 | 14.8 | 7,806 | 11,169 | 34.9 |
| L-5502 | 13,285,841 | 14.0 | 8,623 | 11,364 | 37.2 |
| L-4474 | 12,138,526 | 14.9 | 6,261 | 8,928 | 24.6 |
| L-7657 | 5,499,772 | 14.9 | 8,946 | 13,083 | 15.9 |
| L-8302 | 12,172,065 | 18.6 | 6,158 | 31,841 | 24.2 |
| L-23100 | 3,996,404 | 12.7 | 22,496 | 36,146 | 29.4 |
| L-24764 | 3,948,551 | 12.2 | 25,424 | 34,084 | 32.9 |
| L-3041 | 5,328,679 | 21.0 | 18,055 | 38,111 | 31.2 |
| L-3049 | 6,320,838 | 20.7 | 16,191 | 37,510 | 33.2 |
| L-7759 | 2,970,312 | 12.6 | 20,034 | 27,913 | 19.4 |
| L-5964 | 6,065,873 | 20.8 | 16,036 | 35,154 | 31.6 |
| 3007-10 | 8,980,110 | 20.3 | 8,649 | 27,681 | 25.1 |
| 3007-14 | 3,155,759 | 12.1 | 23,218 | 34,893 | 24.0 |
| 3048-28 | 2,396,810 | 11.8 | 21,850 | 34,756 | 17.1 |
| 3049-10 | 3,448,353 | 11.8 | 19,595 | 34,271 | 22.1 |
| 3049-06 | 3,138,868 | 12.0 | 22,998 | 35,827 | 23.6 |
| **Mean** | 7,239,301 | 15.3 | 14,587 | 25,805 | 26.8 |
| **Median** | 6,065,873 | 14.9 | 16,036 | 31,841 | 24.6 |
| **SD** | 3,949,356 | 3.3 | 6,936 | 11,441 | 6.3 |

ID: identifier of the sample e.g. L-number, kb: kilobase, Mb: megabase, Q-score: read quality (Phred).

**Supplementary Table 6: Quantity of OGM variants.** All high-quality SVs and CNV identified with optical genome mapping (OGM) and filtered subsets for rare variants and variants in known genes associated with movement disorders are shown.

| **Bionano optical genome mapping** | | | | | |
|---|---|---|---|---|---|
| ID | All SVs, CNVs (unfiltered) | All SVs, CNVs (high QC) | SVs or CNVs in MDS genes | Rare SVs or CNVs | Rare SVs or CNVs in MDS genes |
| L-4035 | 3,899 | 2,696 | 19 | 33 | |
| L-3047 | 4,167 | 2,677 | 20 | 47 | 1 |
| L-3046 | 3,927 | 2,671 | 13 | 41 | |
| L-3050 | 3,755 | 2,583 | 12 | 35 | |
| L-5502 | 3,814 | 1,566 | 9 | 13 | |
| L-4474 | 4,074 | 2,630 | 18 | 55 | |
| L-7657 | 4,073 | 2,618 | 14 | 38 | |
| L-8302 | 3,830 | 2,593 | 11 | 42 | |
| L-23100 | 3,981 | 2,593 | 16 | 29 | |
| L-24764 | 4,135 | 2,695 | 14 | 33 | |
| L-3041 | 4,089 | 2,657 | 14 | 32 | |
| L-3049 | 4,076 | 2,653 | 15 | 37 | 1 |
| L-7759 | 4,293 | 2,842 | 20 | 51 | |
| L-5964 | 4,077 | 2,645 | 16 | 31 | 1 |
| 3007-10 | 4,143 | 2,687 | 15 | 50 | |
| 3007-14 | 4,037 | 2,581 | 16 | 53 | |
| 3048-28 | 4,243 | 2,715 | 20 | 59 | 1 |
| 3049-10 | 4,295 | 2,810 | 19 | 62 | |
| 3049-06 | 4,211 | 2,765 | 18 | 63 | |
| **Sum** | 77,119 | 49,677 | 299 | 804 | 4 |
| **Mean** | 4,056 | 2,597 | 15 | 40 | *NA* |

Variants were considered high quality with Bionano Confidence >0.99 (corresponding to a Phred quality score of 20) and used for further filtering. ID: identifier of the sample, rare: AF ≤0.01 in the internal Bionano control database.

**Supplementary Table 7: Quantity of ONT variants.** All SVs and CNV identified with Nanopore long-read sequencing (ONT) and filtered subsets for rare variants and variants in known genes associated with movement disorders are shown.

| **Oxford Nanopore long-read sequencing** | | | | | |
|---|---|---|---|---|---|
| ID | All SVs, CNVs | All SVs, CNVs (≥500 bp) | SVs or CNVs in MDS genes | Rare SVs or CNVs | Rare SVs or CNVs in MDS genes |
| L-4035 | 20,767 | 4,214 | 83 | 1,285 | 2 |
| L-3047 | 21,676 | 4,874 | 93 | 1,372 | 5 |
| L-3046 | 20,781 | 4,387 | 91 | 1,314 | 4 |
| L-3050 | 21,366 | 4,535 | 95 | 1,320 | 6 |
| L-5502 | 21,437 | 4,689 | 92 | 1,282 | 3 |
| L-4474 | 20,693 | 4,203 | 90 | 1,335 | 5 |
| L-7657 | 21,034 | 4,473 | 89 | 1,262 | 4 |
| L-8302 | 22,569 | 5,041 | 97 | 1,209 | 7 |
| L-23100 | 21,526 | 4,932 | 90 | 1,275 | 5 |
| L-24764 | 21,720 | 4,992 | 89 | 1,361 | 5 |
| L-3041 | 22,983 | 5,287 | 100 | 1,211 | 7 |
| L-3049 | 23,092 | 5,343 | 94 | 1,199 | 5 |
| L-7759 | 23,011 | 5,333 | 92 | 1,482 | 4 |
| L-5964 | 20,851 | 4,755 | 87 | 1,244 | 1 |
| 3007-10 | 23,013 | 5,045 | 95 | 1,248 | 3 |
| 3007-14 | 21,632 | 4,766 | 90 | 1,379 | 3 |
| 3048-28 | 22,165 | 5,119 | 105 | 1,444 | 5 |
| 3049-10 | 22,288 | 5,056 | 92 | 1,487 | 5 |
| 3049-06 | 22,084 | 4,986 | 100 | 1,363 | 5 |
| **Sum** | 414,688 | 92,030 | 1,764 | 25,072 | 84 |
| **Mean** | 21,826 | 4,844 | 93 | 1320 | 4 |

All variants passed the quality criteria (FILTER=PASS) and exhibited a Phred quality score ≥20. Variants with an allele frequency of ≤0.01 in the gnomAD 4.1 SV database were considered rare. ID: identifier of the sample, rare: AF ≤0.01 in gnomAD 4.1 SV.

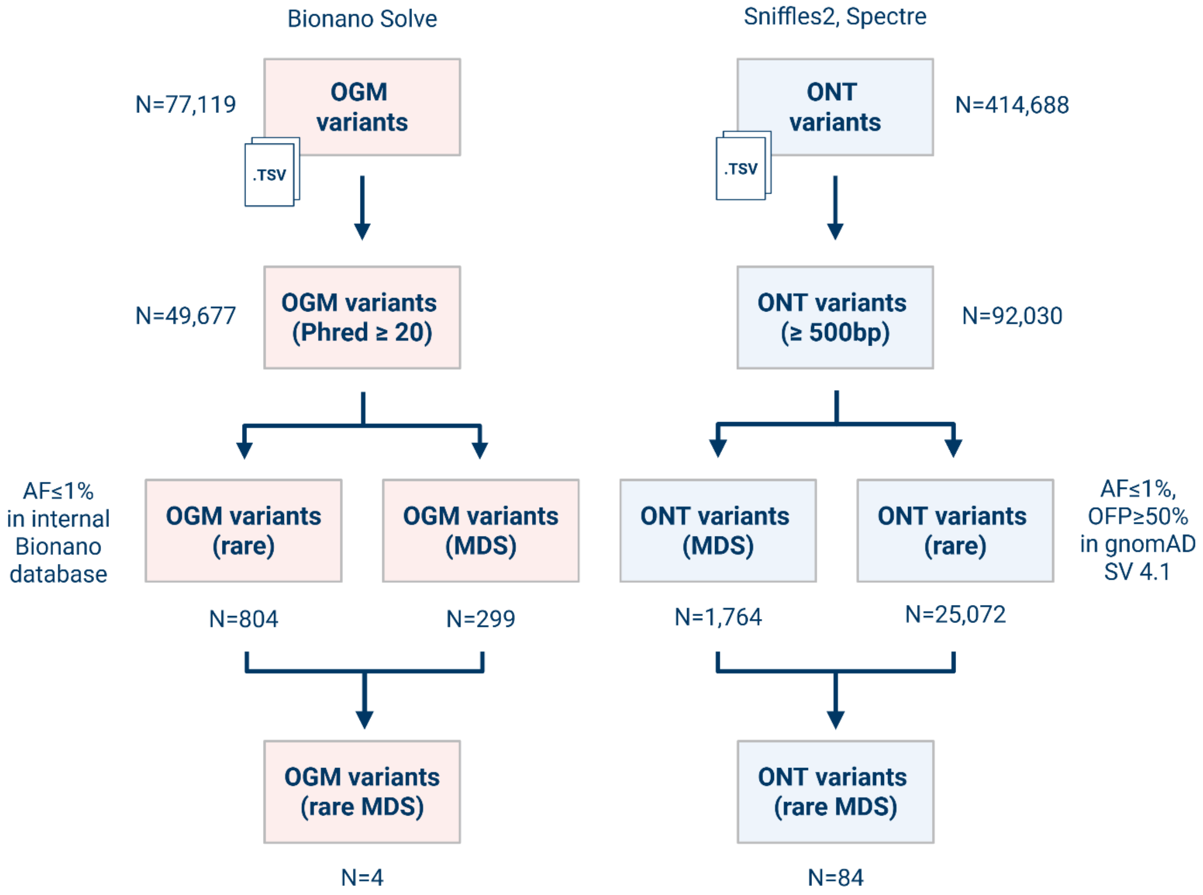


**Supplementary Figure 1: Variant filter steps.** High-quality SVs and CNVs identified with OGM and ONT were filtered based on allele frequency and position within known genes associated with movement disorders. Variants were considered high-quality with a Bionano confidence score greater than 0.99 (corresponding to a Phred quality score of 20). All ONT variants passed the quality criteria (FILTER=PASS) and exhibited a Phred quality score of greater than 20. Variants with an allele frequency of ≤0.01 in the internal Bionano control database (OGM) or the gnomAD 4.1 SV database (ONT) were considered rare. Created in BioRender. Fienemann, A. (2025).

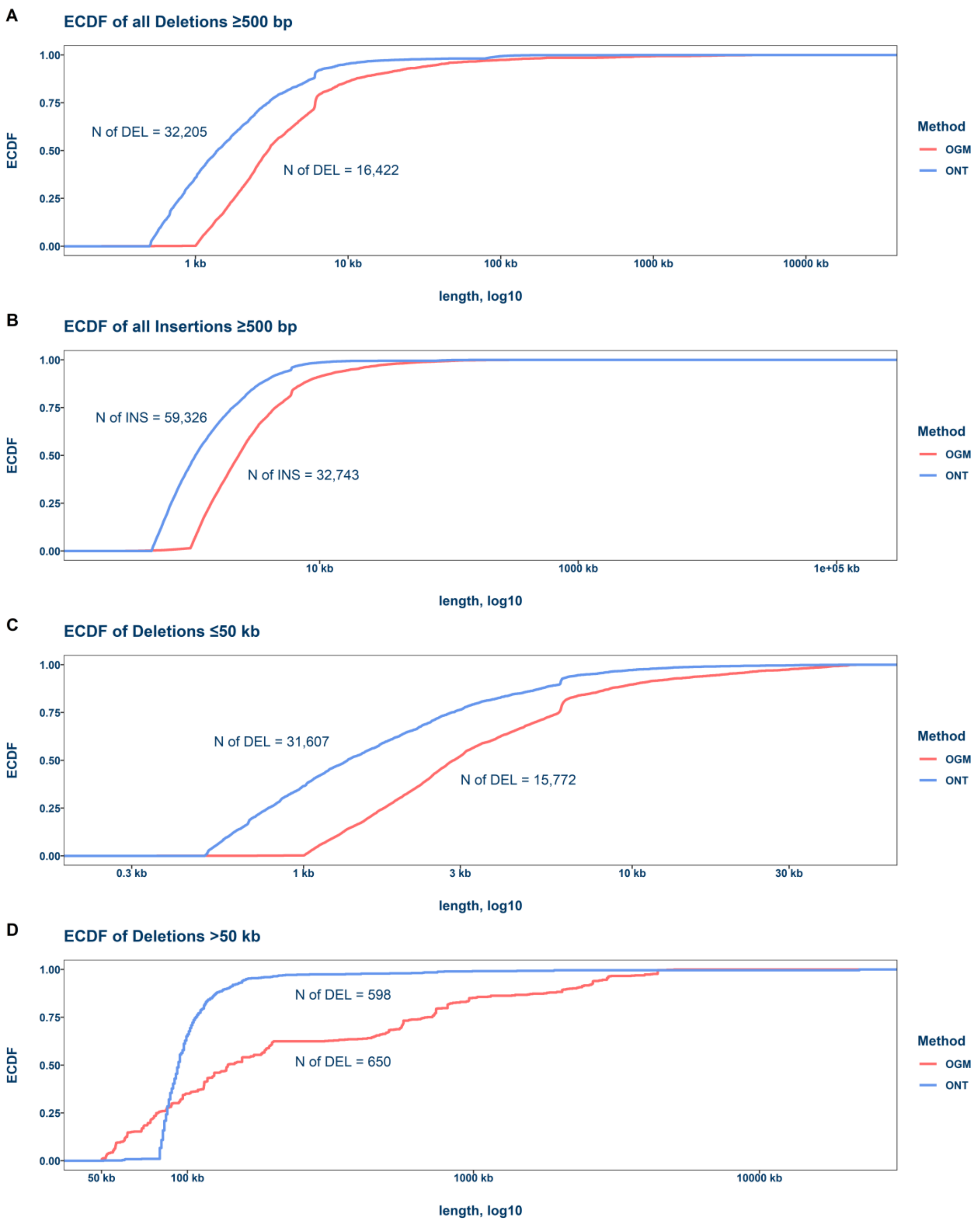


**Supplementary Figure 2: ECDF comparison.** Empirical cumulative distribution functions (ECDFs) illustrate the composition of insertion and deletion lengths detected by OGM and ONT. All OGM ECDFs are shifted to a longer variant length compared to ONT. **A**. Comparison of all deletions over the size of 500 bp (resolution of OGM). **B**. Comparison of all insertions over the size of 500 bp. **C**. Comparison of all deletions ≤50 kb. **D**. Comparison of all deletions >50 kb. INS: insertion, DEL: deletion.

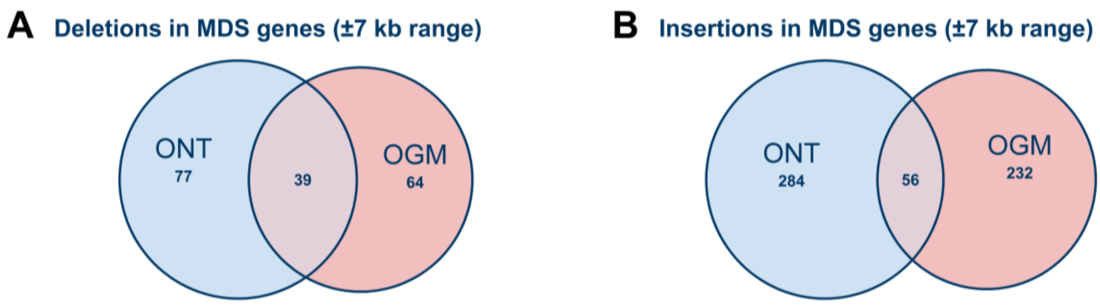


**Supplementary Figure 3: Concordance of MDS variants between ONT and OGM**. These Venn diagrams show the simplified overlap of deletions (**A**) and insertions (**B**) between SVs and CNVs (>500 bp) overlapping with known genes associated with movement disorders identified by ONT and OGM. Due to the difference in methodology, the SVs in the overlap fraction are not exact matches. For the overlap, a fixed interval (±7 kb) regarding the start and end of the variants was applied based on the median 95% confidence interval of all OGM calls affecting MDS genes.

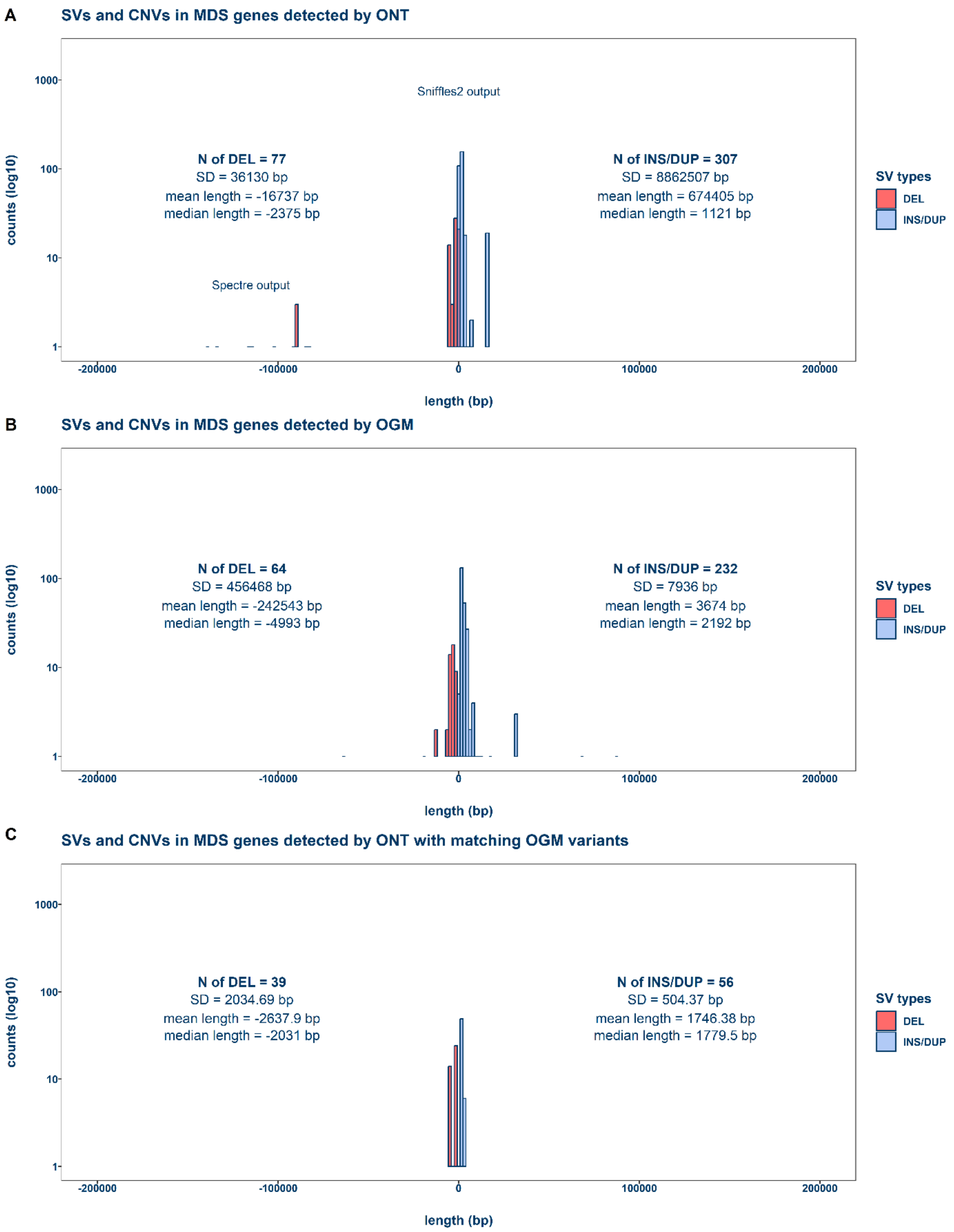


**Supplementary Figure 4: Length distribution of SVs and CNVs in MDS genes**. The mirrored histograms show the length distribution of deletions and insertions/duplications ≥500 bp that overlapped with known genes associated with movement disorders called by ONT and OGM. The variant length in base pairs (bp) is plotted against the logarithmic counts of the variants. **A**: SVs and CNVs in MDS genes identified by Sniffles2 and Spectre in the ONT data. **B**: SVs and CNVs in MDS genes identified by Bionano Solve in the OGM data. **C**: Variants called by ONT with overlapping calls by OGM. DEL: deletion, DUP: duplication, INS: insertion.

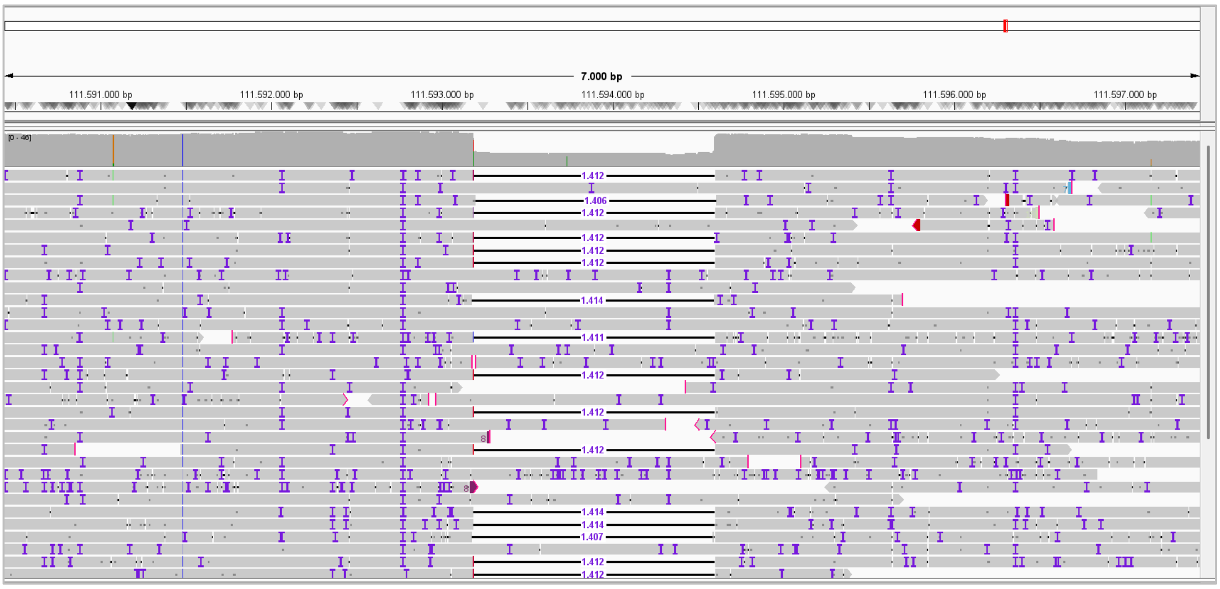


**Supplementary Figure 5: IGV snapshot of a *ATXN2* deletion.** The figure shows a snapshot from the Integrative Genomics Viewer (IGV) displaying ONT reads mapped to Chromosome 12 of the GRCh38 reference genome. A heterozygous 1.4 kb deletion was identified at the position chr12:111593192—111594604.

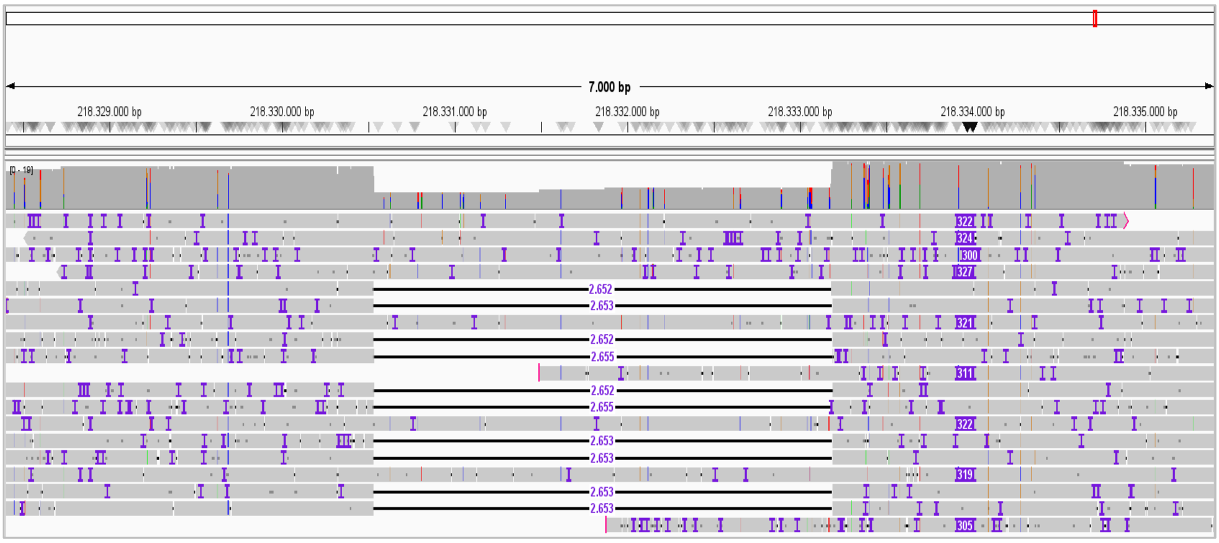


**Supplementary Figure 6: IGV snapshot of a *PNKD* deletion.** The figure shows a snapshot from the Integrative Genomics Viewer (IGV) displaying ONT reads mapped to Chromosome 2 of the GRCh38 reference genome. A heterozygous 2.6 kb deletion was identified at the position chr2:218330537—218333189.

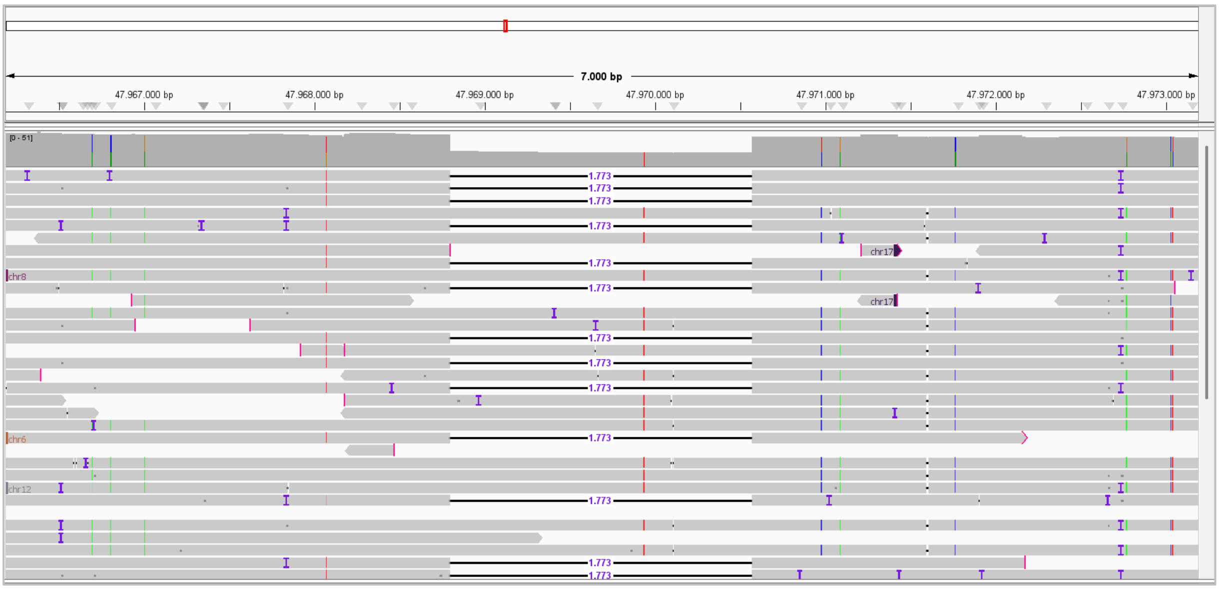


**Supplementary Figure 7: IGV snapshot of a *SUCLA2* deletion.** The figure shows a snapshot from the Integrative Genomics Viewer (IGV) displaying ONT reads mapped to Chromosome 13 of the GRCh38 reference genome. A heterozygous 1.7 kb deletion was identified at the position chr13:47968800—47970573.

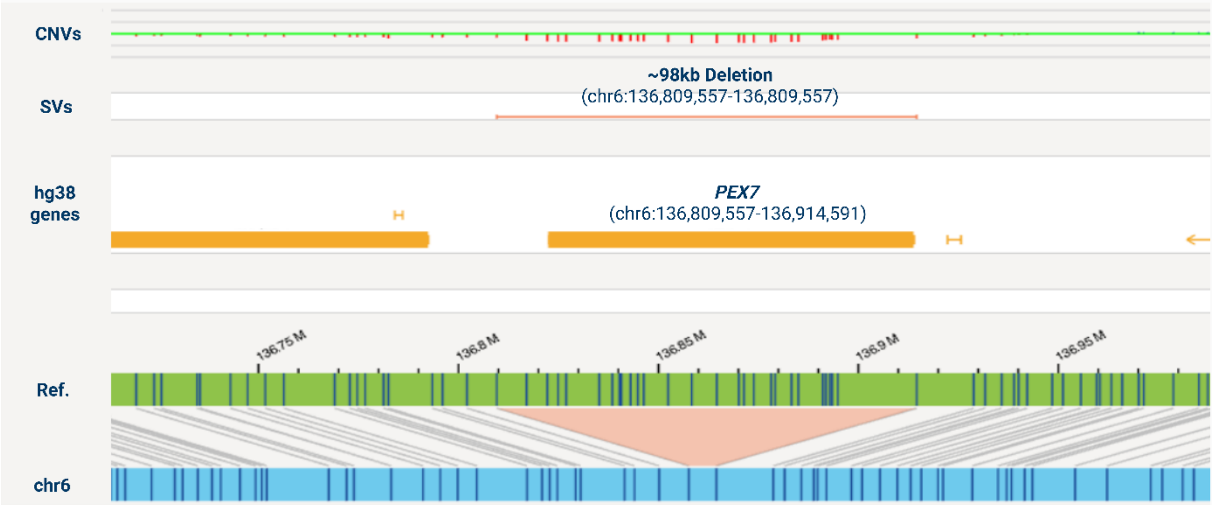


**Supplementary Figure 8: Bionano Access snapshot of a 98 kb deletion spanning *PEX7*.** The figure shows a 98 kb deletion spanning most of the *PEX7* gene. Created in BioRender. Fienemann, A. (2025).

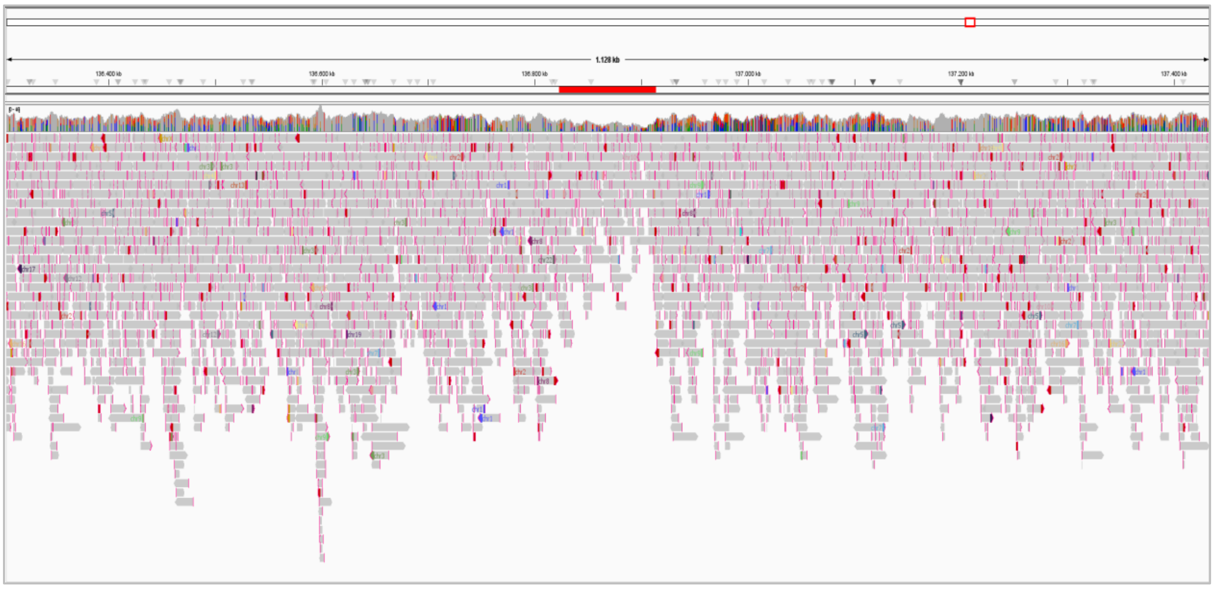

**Supplementary Figure 9: IGV snapshot of a 98kb deletion spanning *PEX7*.** The figure shows a snapshot from the Integrative Genomics Viewer (IGV) displaying ONT reads mapped to Chromosome 6 of the GRCh38 reference genome. A heterozygous copy number reduction affecting *PEX7* (red bar) can be identified visually, indicating a deletion; however, no variant was called with the SV-caller Sniffles2 (v2.3.3).

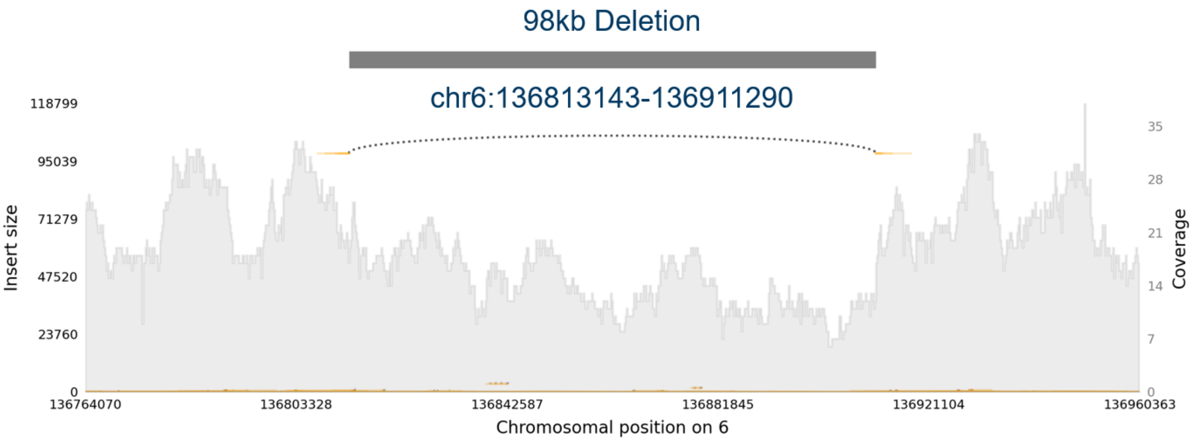


**Supplementary Figure 10: Samplot snapshot of a 98 kb deletion affecting *PEX7*.** The deletion spanning the *PEX7* gene can be detected using v2.6.2 of the SV-caller Sniffles2. The visualization tool Samplot was used (Belyeu et al., 2021).
